# Assessing the causal role of epigenetic clocks in the development of multiple cancers: a Mendelian randomization study

**DOI:** 10.1101/2021.11.29.21266984

**Authors:** Fernanda Morales-Berstein, Daniel L McCartney, Ake T Lu, Konstantinos K Tsilidis, Emmanouil Bouras, Philip Haycock, Kimberley Burrows, Amanda I Phipps, Daniel D Buchanan, Iona Cheng, the PRACTICAL consortium, Richard M Martin, George Davey Smith, Caroline L Relton, Steve Horvath, Riccardo E Marioni, Tom G Richardson, Rebecca C Richmond

## Abstract

**Background:** Epigenetic clocks have been associated with cancer risk in several observational studies. Nevertheless, it is unclear whether they play a causal role in cancer risk or if they act as a non-causal biomarker.

**Methods:** We conducted a two-sample Mendelian randomization (MR) study to examine the genetically predicted effects of epigenetic age acceleration as measured by HannumAge (9 single-nucleotide polymorphisms (SNPs)), Horvath Intrinsic Age (24 SNPs), PhenoAge (11 SNPs) and GrimAge (4 SNPs) on multiple cancers (i.e., breast, prostate, colorectal, ovarian and lung cancer). We obtained genome-wide association data for biological ageing from a meta-analysis (N=34,710), and for cancer from the UK Biobank (N cases=2,671–13,879; N controls=173,493–372,016), FinnGen (N cases=719–8,401; N controls=74,685–174,006) and several international cancer genetic consortia (N cases=11,348–122,977; N controls=15,861–105,974). Main analyses were performed using multiplicative random effects inverse variance weighted (IVW) MR. Individual study estimates were pooled using fixed effect meta-analysis. Sensitivity analyses included MR-Egger, weighted median, weighted mode and Causal Analysis using Summary Effect Estimates (CAUSE) methods, which are robust to some of the assumptions of the IVW approach.

**Results:** Meta-analysed IVW MR findings suggested that higher GrimAge acceleration increased the risk of colorectal cancer (OR=1.12 per year increase in GrimAge acceleration, 95%CI 1.04–1.20, *p*=0.002). The direction of the genetically predicted effects was consistent across main and sensitivity MR analyses. Among subtypes, the genetically predicted effect of GrimAge acceleration was greater for colon cancer (IVW OR=1.15, 95%CI 1.09–1.21, *p*=0.006), than rectal cancer (IVW OR=1.05, 95%CI 0.97–1.13, *p*=0.24). We also found evidence that higher GrimAge acceleration decreased the risk of prostate cancer (pooled IVW OR=0.93 per year increase in GrimAge acceleration, 95%CI 0.87–0.99, *p*=0.02). This was supported by MR sensitivity analyses, but did not replicate in MR analyses using data on parental history of prostate cancer in UK Biobank (IVW OR=1.00, 95%CI 0.96–1.04, *p*=1.00). Results were less consistent for associations between other epigenetic clocks and cancers.

**Conclusions:** GrimAge acceleration may increase the risk of colorectal cancer. Additionally, there is more limited evidence that it may be protective against prostate cancer. Findings for other clocks and cancers were inconsistent. Further work is required to investigate the potential mechanisms underlying the results.

**Funding:** FMB was supported by a Wellcome Trust PhD studentship in Molecular, Genetic and Lifecourse Epidemiology (218495/Z/19/Z). KKT was supported by a Cancer Research UK (C18281/A29019) programme grant (the Integrative Cancer Epidemiology Programme) and by the Hellenic Republic’s Operational Programme “Competitiveness, Entrepreneurship & Innovation” (OΠΣ 5047228). PH was supported by Cancer Research UK (C18281/A29019).

RMM was supported by the NIHR Biomedical Research Centre at University Hospitals Bristol and Weston NHS Foundation Trust and the University of Bristol and by a Cancer Research UK (C18281/A29019) programme grant (the Integrative Cancer Epidemiology Programme). The views expressed are those of the author(s) and not necessarily those of the NIHR or the Department of Health and Social Care. GDS and CLR were supported by the Medical Research Council (MC_UU_00011/1 and MC_UU_00011/5) and by a Cancer Research UK (C18281/A29019) programme grant (the Integrative Cancer Epidemiology Programme). REM was supported by an Alzheimer’s Society project grant (AS-PG-19b-010) and NIH grant (U01 AG-18-018, PI: Steve Horvath). RCR is a de Pass Vice Chancellor’s Research Fellow at the University of Bristol.

## INTRODUCTION

DNA methylation (DNAm) is an epigenetic biomarker that can be used as an estimator of chronological age. Biological age, as predicted by DNAm patterns at specific cytosine-phosphate-guanine (CpG) sites, may differ from chronological age on an individual basis. Observational evidence suggests that epigenetic age acceleration (i.e., when an individual’s biological age is greater than their chronological age) may be associated with an increased risk of mortality and age-related diseases, including cancer^1^.

Epigenetic clocks are heritable indicators of biological ageing derived from DNAm data. Each clock is based on DNAm levels measured at a different set of CpG sites, which capture distinctive features of epigenetic ageing^2^. ‘First-generation’ epigenetic clocks, such as HannumAge^3^ and Intrinsic HorvathAge^4^, have been derived from DNAm levels at CpG sites found to be strongly associated with chronological age. HannumAge is trained on 71 age-related CpGs found in blood^3^, while Intrinsic HorvathAge is based on 353 age-related CpGs found in several human tissues and cell types, and is further adjusted for blood cell counts^4^. More recently, ‘second-generation’ epigenetic clocks, such as, PhenoAge^5^ and GrimAge^6^, have been developed to predict age-related morbidity and mortality. PhenoAge incorporates data from 513 age-related CpGs associated with mortality and nine clinical biomarkers (i.e., albumin, creatinine, serum glucose, C-reactive protein, lymphocyte percentage, mean corpuscular volume, red cell distribution width, alkaline phosphatase and leukocyte count)^5^, and GrimAge includes data from 1,030 age-related CpGs associated with smoking pack-years and seven plasma proteins (i.e., cystatin C, leptin, tissue inhibitor metalloproteinases 1, adrenomedullin, beta-2-microglobulin, growth differentiation factor 15, and plasminogen activation inhibitor 1 (PAI-1))^6^. Due to differences in their composition, HannumAge and Intrinsic HorvathAge are better predictors of chronological age^3^ ^4^, while PhenoAge and GrimAge stand out for their ability to predict health and lifespan^5–7^.

Several studies suggest that HannumAge, Intrinsic HorvathAge, PhenoAge and GrimAge acceleration are associated with cancer risk^5^ ^8–13^. In contrast, others indicate that evidence in support of this claim is weak or non existent^14–17^. This lack of consensus could be explained by biases that often affect observational research, such as reverse causation (e.g., cancer influencing the epigenome and not the other way around) and residual confounding (e.g., unmeasured, or imprecisely measured confounders of the association between epigenetic age acceleration and cancer)^18^.

The strength of the associations between epigenetic age acceleration and different cancers has also been found to vary across epigenetic clocks. For instance, positive associations between epigenetic age acceleration and colorectal cancer seem to be much stronger when biological age is estimated using second-generation clocks (i.e., PhenoAge and GrimAge)^10^ rather than first-generation clocks (i.e., HannumAge and Intrinsic HorvathAge)^14^ ^16^. Lack of consensus across epigenetic clocks could be explained by differences in their algorithms (which may reflect different mechanisms of biological ageing), as well as heterogeneity in study designs^1^. Furthermore, even if there were a consensus, it would still be unclear whether age-related DNA methylation plays a causal role in cancer risk or if it merely acts as a non-causal prognostic biomarker.

Mendelian randomization (MR), a method that uses genetic variants as instrumental variables to infer causality between a modifiable exposure and an outcome, is less likely to be affected by residual confounding and reverse causation than traditional observational methods^19^. A recent genome-wide association study (GWAS) meta-analysis has revealed 137 genetic loci associated with epigenetic age acceleration (as measured by six epigenetic biomarkers) that may be used within an MR framework^20^.

McCartney et al.^20^ used IVW MR, MR-Egger, weighted median and weighted mode methods to explore the genetically predicted effects of HannumAge, Intrinsic HorvathAge, PhenoAge and GrimAge acceleration on breast, ovarian and lung cancer. Here we extend this analysis to include colorectal and prostate cancer (two of the most common cancers worldwide^21^) and use additional methods and datasets to verify the robustness of our findings.

The aim of this two-sample MR study was to examine the genetically predicted effects of epigenetic age acceleration (as measured by HannumAge^3^, Horvath Intrinsic Age^4^, PhenoAge^5^ and GrimAge^6^) on multiple cancers (i.e., breast, prostate, colorectal, ovarian and lung cancer) using summary genetic association data from (1) McCartney et al. (N=34,710)^20^, (2) the UK Biobank (N cases=2,671–13,879; N controls=173,493–372,016), (3) FinnGen (N cases=719– 8,401; N controls=74,685–174,006) and (4) several international cancer genetic consortia (N cases=11,348–122,977; N controls=15,861–105,974).

## METHODS

### Reporting guidelines

This study has been reported according to the STROBE-MR guidelines^22^ (**Supplementary material**).

### Genetic instruments for epigenetic age acceleration

We obtained summary genetic association estimates for epigenetic age acceleration measures of HannumAge^3^, Intrinsic HorvathAge^4^, PhenoAge^5^ and GrimAge^6^ from a recent GWAS meta-analysis of biological ageing^20^, which included 34,710 participants of European ancestry. Across the 28 European ancestry studies considered in the analysis, 57.3% of participants were female. A detailed description of the methods that were used can be found in the publication^20^. In short, outlier samples with clock methylation estimates of +/− 5 s.d. from the mean were excluded from further analysis. SNPs were genotyped and imputed independently for each cohort included in the meta-analysis and genotypes were imputed using either the HRC or the 1000 Genomes Project Phase 3 reference panels.

We selected GWAS-significant SNPs (*p*<5×10^−8^) for each epigenetic age acceleration measure and performed linkage disequilibrium (LD) clumping (r^2^<0.001) to remove correlated variants using the European reference panel from the 1000 Genomes Project Phase 3 v5.

We identified 9 independent SNPs for HannumAge, 24 for Intrinsic HorvathAge, 11 for PhenoAge and 4 for GrimAge (**Supplementary Table 1**). The genetic instruments for HannumAge, Intrinsic HorvathAge, PhenoAge and GrimAge acceleration explained 1.48%, 4.41%, 1.86% and 0.47% of the trait variance, respectively. All the selected SNPs had F-statistics greater than 10 (HannumAge median 38 and range 31–99, Intrinsic HorvathAge median 47 and range 31–240, PhenoAge median 45 and range 32–89, GrimAge median 36 and range 31–45).

### Genetic association data sources for cancer outcomes

We obtained summary-level genetic association data for cancer outcomes from the UK Biobank and several international cancer genetic consortia (**Table 1**). Data were obtained from UK Biobank, FinnGen, the Breast Cancer Association Consortium (BCAC), the Ovarian Cancer Association Consortium (OCAC), the Consortium of Investigators of Modifiers of BRCA1/2 (CIMBA), the Prostate Cancer Association Group to Investigate Cancer Associated Alterations in the Genome (PRACTICAL), the International Lung Cancer Consortium (ILCCO) and the Genetics and Epidemiology of Colorectal Cancer Consortium (GECCO). Further details of the studies and the data obtained are described in the ***Supplementary Methods***.

**Table 1.** Numbers of overall cancer cases and controls by data source.

| Cancer type | Source | N cases (%) * | N controls |
| --- | --- | --- | --- |
| Breast | BCAC | 122,977 (53.7%) | 105,974 |
|  | UK Biobank | 13,879 (6.5%) | 198,523 |
|  | FinnGen | 8,401 (7.8%) | 99,321 |
| Ovarian | OCAC | 25,509 (38.4%) | 40,941 |
|  | UK Biobank | 1,218 (0.6%) | 198,523 |
|  | FinnGen | 719 (0.7%) | 99,321 |
| Prostate | PRACTICAL | 79,148 (56.4%) | 61,106 |
|  | UK Biobank | 9,132 (5.0%) | 173,493 |
|  | FinnGen | 6,311 (7.8%) | 74,685 |
| Lung | ILCCO | 11,348 (41.7%) | 15,861 |
|  | UK Biobank | 2,671(0.7%) | 372,016 |
|  | FinnGen | 1,681 (1.0%) | 173,933 |
| Colorectal | GECCO | 58,131 (46.3%) | 67,347 |
|  | UK Biobank | 5,657 (1.5%) | 372,016 |
|  | FinnGen | 3,022 (1.7%) | 174,006 |

Abbreviations: BCAC, Breast Cancer Association Consortium; OCAC, Ovarian Cancer Association Consortium; PRACTICAL, Prostate Cancer Association Group to Investigate Cancer Associated Alterations in the Genome; ILCCO, International Lung Cancer Consortium; GECCO, Genetics and Epidemiology of Colorectal Cancer Consortium.
\*Percentage (%) of cases within each study source was calculated using the following formula: $100 * N \text{ cases} / (N \text{ cases} + N \text{ controls})$

We extracted genetic association data for the selected SNPs from each cancer GWAS (for breast, prostate, colorectal, ovarian and lung cancers). LD proxies (r^2^>0.8) were used when the SNPs of interest were missing from the cancer GWAS dataset. The proxies were located using the MR-Base platform, which calculates LD using the European subset of individuals from the 1000 Genomes Project reference panel as above^23^. The “LDlinkR” R package version 1.1.2 was used to find proxies for cancer data that were not included in the MR-Base platform. The exposure and outcome datasets were then harmonised to ensure the genetic associations reflect the same effect allele. Palindromic SNPs with minor allele frequencies (MAF) <0.3 were aligned, while those with MAF ≥0.3 or mismatching strands were excluded.

### Power calculations

Statistical power was calculated using an online calculator for MR available at: https://shiny.cnsgenomics.com/mRnd/. Calculations were performed separately for each clock-cancer combination. They were based on a type 1 error rate of 0.05, the proportion of phenotypic variance explained by genetic variants (R^2^) for each measure of epigenetic age acceleration, and the total number of cases and controls included in the meta-analysis for each cancer. Across combinations of the four epigenetic clock acceleration and five cancer measures, we had 80% power to detect ORs as small as 1.04–1.39 (**Supplementary Table 2**).

### Statistical analysis

We estimated the genetically predicted effects of epigenetic age acceleration (as measured by HannumAge^3^, Horvath Intrinsic Age^4^, PhenoAge^5^ and GrimAge^6^) on multiple cancers (i.e., breast, prostate, colorectal, ovarian and lung cancer) using a two-sample MR framework (**Figure 1**).

**Figure 1.**
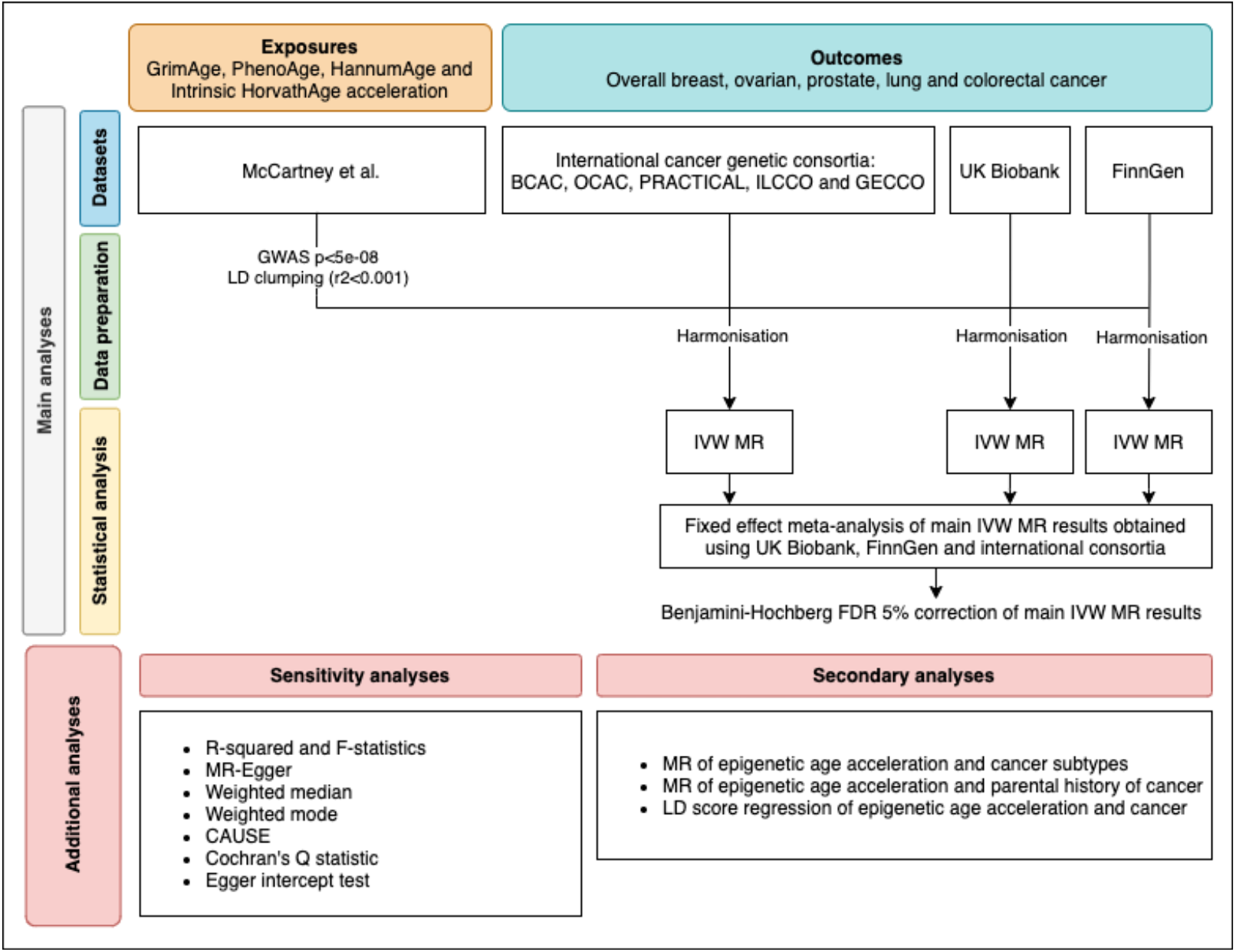
Flowchart summarising study methods. Abbreviations: BCAC, Breast Cancer Association Consortium; OCAC, Ovarian Cancer Association Consortium; PRACTICAL, Prostate Cancer Association Group to Investigate Cancer Associated Alterations in the Genome; ILCCO, International Lung Cancer Consortium; GECCO, Genetics and Epidemiology of Colorectal Cancer Consortium; LD, linkage disequilibrium; IVW, inverse variance weighted; MR, Mendelian randomization; FDR, false discovery rate; GWAS, genome-wide association study; CAUSE, Causal Analysis Using Summary Effect estimates.

#### Main analyses

Main analyses were performed using multiplicative random effects inverse variance weighted (IVW) MR, a method that combines the genetically predicted effect of epigenetic age acceleration on cancer across genetic variants^24^. We used fixed effect meta-analysis to pool results across studies (i.e., UK Biobank, FinnGen and international consortia). For colorectal cancer, we only pooled FinnGen and GECCO estimates, since UK Biobank participants were already included in GECCO. I^2^ statistics and their corresponding confidence intervals were used to estimate heterogeneity across study estimates^25^. A Benjamini-Hochberg false discovery rate (FDR) <5% was used to correct the pooled main IVW results for multiple testing^26^.

#### Sensitivity analyses

MR assumes genetic instruments for epigenetic age acceleration are 1) associated with epigenetic age acceleration (relevance assumption), 2) independent of confounders (independence assumption) and 3) only associated with cancer through their effect on epigenetic age acceleration (exclusion restriction assumption)^27^ ^28^.

As a sensitivity analysis and to test for potential violations of the relevance assumption, we calculated F-statistics and the R^2^ for each measure of epigenetic age acceleration^29^. Other sensitivity analyses included MR-Egger^30^, weighted median^31^ and weighted mode^32^ methods, which are robust to some of the assumptions of the IVW approach (described in the ***Supplementary Methods***). These results were also pooled across studies, as explained above. Consistency across different MR methods would suggest that it is less likely that the independence and exclusion restriction assumptions are violated.

Where associations between genetically predicted epigenetic age acceleration and cancer were identified, we additionally performed single-SNP two-sample MR analysis to assess whether the effects were likely to be driven by a single SNP. We also used Causal Analysis using Summary Effect Estimates (CAUSE)^33^, a method that uses genome-wide summary statistics to disentangle causality (i.e., SNPs are associated with cancer through their effect on epigenetic age acceleration) from correlated horizontal pleiotropy (i.e., SNPs are associated with epigenetic age acceleration and cancer through a shared heritable factor), while taking into account uncorrelated horizontal pleiotropy (i.e., SNPs are associated with epigenetic age acceleration through separate mechanisms). It uses Bayesian modelling to assess whether the sharing model (i.e., model that fixes the causal effect at zero) fits the data at least as well as the causal model (i.e., model that allows a causal effect different from zero). Additionally, Cochran’s Q statistics were used to quantify global heterogeneity across SNP-specific MR estimates^34^ and MR-Egger intercept tests were performed to detect uncorrelated horizontal pleiotropy^30^.

#### Secondary analyses

As a secondary analysis, we conducted two-sample MR of epigenetic age acceleration and cancer subtypes (i.e., breast cancer: ER+, ER-, triple negative, luminal B/HER2-negative-like, HER2-enriched-like, luminal A-like, luminal B-like, BRCA1 and BRCA2; ovarian cancer: high grade serous, low grade serous, invasive mucinous, clear cell, endometrioid, BRCA1 and BRCA2; prostate cancer: advanced, advanced [vs non-advanced], early onset, high risk [vs low risk], and high risk [vs low and intermediate risk]; lung cancer: adenocarcinoma and squamous cell; colorectal cancer: colon-specific, proximal colon-specific, distal colon-specific, rectal-specific, male and female) (***Supplementary Methods***).

We also performed two-sample MR analyses of epigenetic age acceleration and parental history of cancer in the UK Biobank for breast, prostate, lung and bowel cancer (***Supplementary Methods***). Data on parental history of ovarian cancer were not available in UK Biobank. Family history data correlate with combined hospital record and questionnaire data and it has been suggested that they provide better power to detect GWAS-significant associations for some phenotypes in the UK Biobank^35^. Therefore, we expected these results to be consistent with those obtained in the main analyses. Although these analyses were conducted as a form of replication, their effect estimates are not directly comparable to those obtained in the main analyses due to cases in the GWAS-by-proxy of parental endpoints being defined as either or both parents reportedly having a type of cancer.

MR results were reported as the odds ratio (OR) of site-specific cancer per one year increase in genetically predicted epigenetic age acceleration.

LD Score regression^36^ was used to identify genome-wide genetic correlations between epigenetic age acceleration and cancer. Genetic correlations were estimated using full GWAS summary statistics for the epigenetic clocks and cancer, as well as the 1000 Genomes Project European LD reference panel. Traits with mean heritability chi-square values <1.02 were excluded from the analyses.

All MR analyses were performed using R software version 4.0.2. Two sample MR analyses were conducted using the “TwoSampleMR” package version 0.5.5. Meta-analyses of IVW results were performed using the “meta” package version 4.18. CAUSE analyses were conducted using the “cause” package version 1.2.0. Forest plots were created using the “ggforestplot” package version 0.1.0. LD Scores were computed using the “ldsc” command line tool version 1.0.1. The code used in this study is available at: https://github.com/fernandam93/epiclocks_cancer.

## RESULTS

### Breast cancer

We did not find strong evidence of causality between epigenetic age acceleration and breast cancer (GrimAge IVW OR=0.98, 95%CI 0.95–1.00, *p*=0.08; PhenoAge IVW OR=0.99, 95%CI 0.98–1.01, *p*=0.23; HannumAge IVW OR=0.99, 95%CI 0.97–1.02, *p*=0.63; and Intrinsic HorvathAge IVW OR=0.99, 95%CI 0.98–1.00, *p*=0.13) (**Figure 2, Supplementary Figures 1 and 5–7, Supplementary Tables 3–6 and 9–10**).

**Figure 2.**
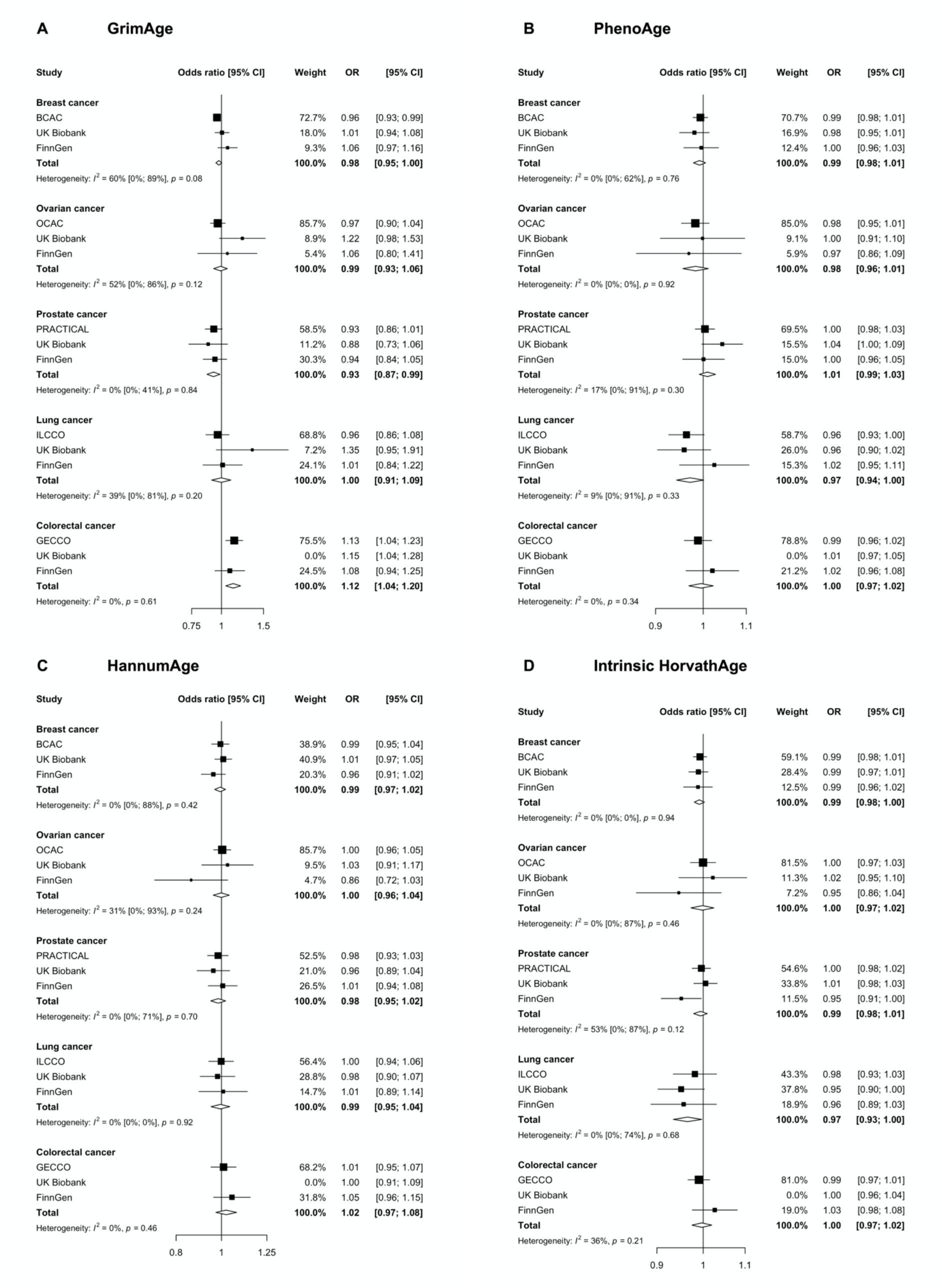
Fixed effect meta-analysis of inverse-variance weighted Mendelian randomization estimates for genetically predicted effects of epigenetic age acceleration on multiple cancers. Odds ratios and 95% confidence intervals are reported per 1 year increase in (A) GrimAge acceleration, (B) PhenoAge acceleration, (C) HannumAge acceleration and (D) Intrinsic HorvathAge acceleration. GrimAge, PhenoAge, HannumAge and Intrinsic HorvathAge acceleration were instrumented by 4, 11, 9 and 24 genetic variants, respectively. All meta-analysis estimates were calculated using data from UK Biobank, FinnGen and international consortia, except for colorectal cancer estimates, which exclude UK Biobank data to avoid double counting.

### Ovarian cancer

There was also limited evidence of causality between epigenetic age acceleration and ovarian cancer (GrimAge IVW OR=0.99, 95%CI 0.93–1.06, *p*=0.78; PhenoAge IVW OR=0.98, 95%CI 0.96–1.01, *p*=0.24; HannumAge IVW OR=1.00, 95%CI 0.96–1.04, *p*=0.95; and Intrinsic HorvathAge IVW OR=1.00, 95%CI 0.97–1.02, *p*=0.89) (**Figure 2, Supplementary Figures 1 and 5–7, Supplementary Tables 3–6 and 9–10**).

### Prostate cancer

Meta-analysed IVW MR findings suggested that genetically predicted GrimAge acceleration decreased the risk of prostate cancer (OR=0.93 per year increase in GrimAge acceleration, 95%CI 0.87–0.99, *p*=0.02) (**Figure 2, Supplementary Tables 3–6**). These results withstood multiple testing correction (FDR *p*=0.04), there was little evidence of heterogeneity across study estimates (I^2^=0%, 95%CI 0–41%, *p*=0.84) and the direction of the genetically predicted effect was consistent across main and sensitivity MR analyses (i.e., MR-Egger, weighted median and weighted mode) (**Figure 3, Supplementary Tables 3–6**).

**Figure 3.**
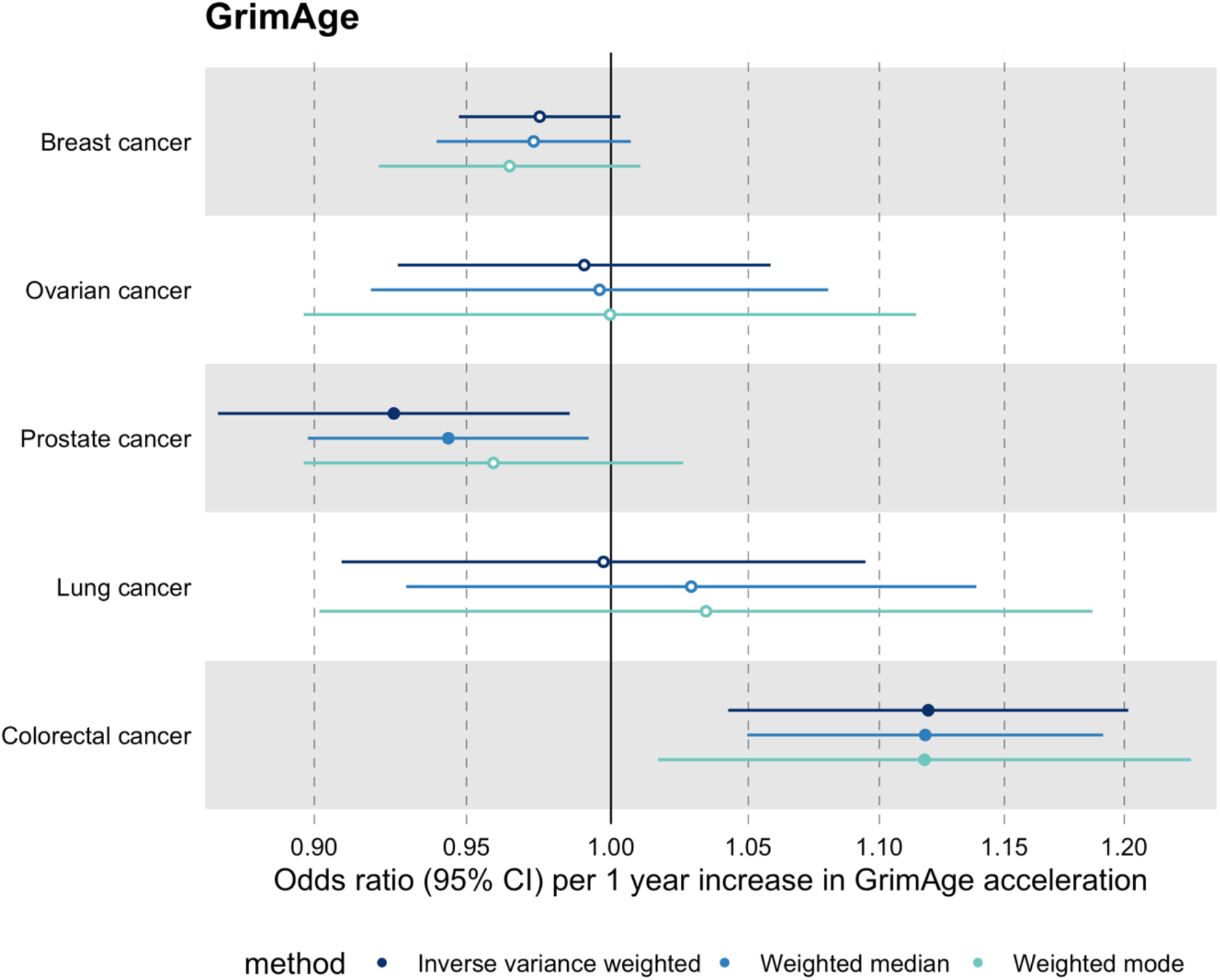
Fixed effect meta-analysis of Mendelian randomization estimates for genetically predicted effects of GrimAge acceleration on multiple cancers. Odds ratios and 95% confidence intervals are reported per 1 year increase in GrimAge acceleration. GrimAge acceleration was instrumented by four genetic variants. Results were obtained using inverse variance weighted MR (dark blue), weighted median (sky blue) and weighted mode (turquoise) methods. All meta-analysis estimates were calculated using data from UK Biobank, FinnGen and international consortia, except for colorectal cancer estimates, which exclude UK Biobank data to avoid double counting.

We further explored the genetically predicted effect of GrimAge on prostate cancer using PRACTICAL data only. Single-SNP analysis revealed that the effect was not driven by a single SNP (**Supplementary Table 7**), although there was some evidence of heterogeneity across SNPs (Cochran’s Q 10.63, *p*= 0.01) (**Supplementary Table 8**). Additionally, results suggested no evidence of detectable uncorrelated horizontal pleiotropy (MR-Egger intercept = -0.03, 95%CI -0.34–0.28, *p*=0.87) (**Supplementary Table 8**). However, we were unable to rule out correlated pleiotropy (CAUSE OR=0.98, 95%CI 0.94–1.02, *p*=0.64; shared *q*=3%, 95%CI=0– 25%) (**Supplementary Figure 2**).

Among subtypes, the protective effect of GrimAge acceleration appeared strongest in relation to advanced (IVW OR=0.90, 95%CI 0.82–1.00, *p*=0.04) and early onset prostate cancer (IVW OR=0.76, 95%CI 0.62–0.92, *p*=0.005) (**Figure 4, Supplementary Table 9**).

**Figure 4.**
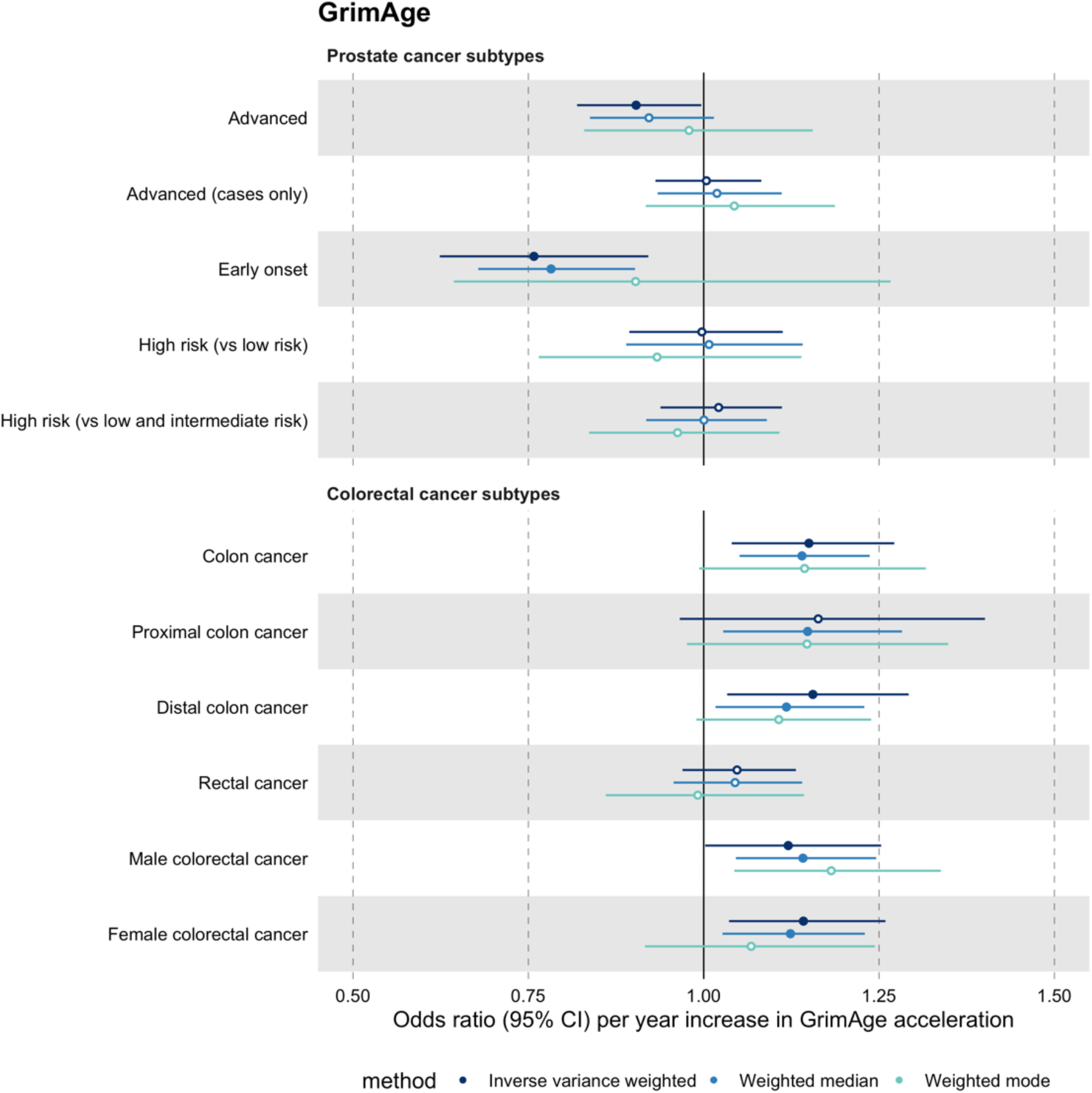
Mendelian randomization estimates for genetically predicted effects of GrimAge acceleration on prostate and colorectal cancer subtypes. Odds ratios and 95% confidence intervals are reported per 1 year increase in GrimAge acceleration. GrimAge acceleration was instrumented by four genetic variants. Results were obtained using inverse variance weighted MR (dark blue), weighted median (sky blue) and weighted mode (turquoise) methods. Data sources: PRACTICAL and GECCO.

However, the prostate cancer results must be interpreted with caution, as findings did not replicate when using parental history of prostate cancer as the outcome (IVW OR=1.00 per year increase in GrimAge acceleration, 95%CI 0.96–1.04, *p*=1.00) (**Figure 5, Supplementary Table 10**). Furthermore, LD Score regression results did not provide strong, consistent evidence of a genetic correlation between GrimAge and prostate cancer (PRACTICAL rg=- 0.13, *p*=0.12; UK Biobank rg=0.03, *p*=0.76; FinnGen rg=-0.02, *p*=0.89) (**Supplementary Figure 3, Supplementary Table 11**)

**Figure 5.**
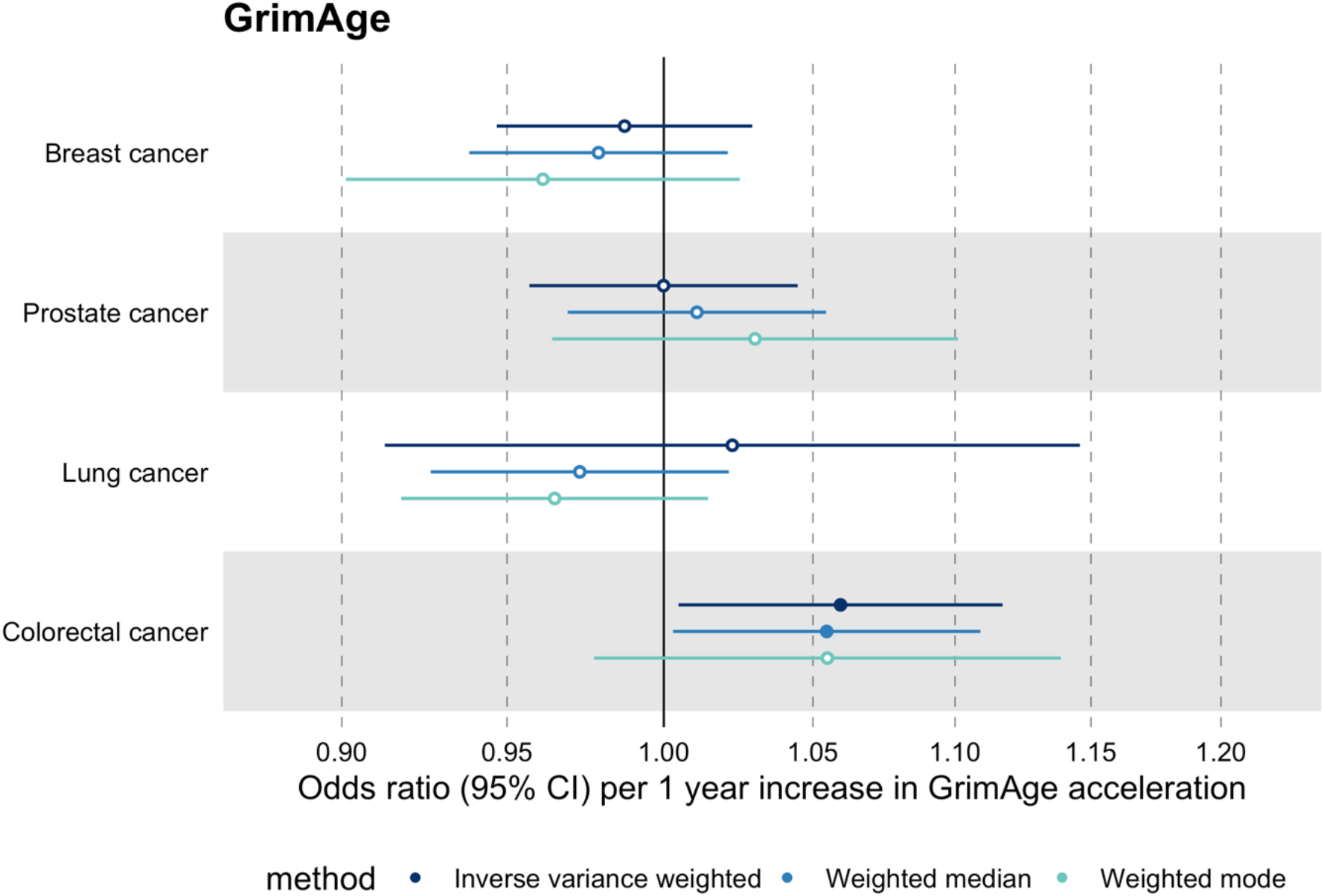
Mendelian randomization estimates for genetically predicted effects of GrimAge acceleration on parental history of multiple cancers. Odds ratios and 95% confidence intervals are reported per 1 year increase in GrimAge acceleration. GrimAge acceleration was instrumented by four genetic variants. Results were obtained using inverse variance weighted MR (dark blue), weighted median (sky blue) and weighted mode (turquoise) methods. Data source: UK Biobank.

We did not find consistent evidence of causality between other measures of epigenetic age acceleration and prostate cancer (PhenoAge IVW OR=1.01, 95%CI 0.99–1.03, *p*=0.31; HannumAge IVW OR=0.98, 95%CI 0.95–1.02, *p*=0.39; and Intrinsic HorvathAge IVW OR=0.99, 95%CI 0.98–1.01, *p*=0.42) (**Figure 2, Supplementary Figures 1 and 5–7, Supplementary Tables 3–6 and 9–10).**

### Lung cancer

Meta-analysed IVW MR findings suggested that genetically predicted Intrinsic HorvathAge acceleration decreased the risk of lung cancer (OR=0.97 per year increase in Intrinsic HorvathAge acceleration, 95%CI 0.93–1.00, *p*=0.03) (**Figure 2, Supplementary Tables 3–6**). However, these results did not survive multiple testing correction (FDR *p*=0.16) and were not strongly supported by sensitivity analyses (**Supplementary Figure 1, Supplementary Tables 3–6**).

We did not find evidence of causality between other measures of epigenetic age acceleration and lung cancer (GrimAge IVW OR=1.00, 95%CI 0.91–1.09, *p*=0.96; PhenoAge IVW OR=0.97, 95%CI 0.94–1.00, *p*=0.06; and HannumAge IVW OR=0.99, 95%CI 0.95–1.04, *p*=0.82) (Figure 2, Supplementary Figures 1 and 5–7, Supplementary Tables 3–6 and 9–10).

### Colorectal cancer

Meta-analysed IVW MR findings suggested that genetically predicted GrimAge acceleration increased the risk of colorectal cancer (OR=1.12 per year increase in GrimAge acceleration, 95%CI 1.04–1.20, *p*=0.002) (**Figure 2, Supplementary Tables 3, 5 and 6**). These results survived multiple testing correction (FDR *p*=0.009) and there was little evidence of heterogeneity across FinnGen and GECCO estimates (I^2^=0%, 95%CI “NA”, *p*=0.61).

Additionally, the direction of the genetically predicted effect was consistent across main and sensitivity MR analyses (i.e., MR-Egger, weighted median and weighted mode) (**Figure 3, Supplementary Tables 3, 5 and 6**) and results replicated when using UK Biobank data alone (IVW OR=1.15, 95%CI 1.04–1.28, *p*=0.007) (**Figure 2, Supplementary Table 4**).

We further explored the genetically predicted effect of GrimAge on colorectal cancer using GECCO data only. Single-SNP analysis revealed that the effect was not driven by a single SNP (**Supplementary Table 7**). Moreover, there was no detectable evidence of uncorrelated horizontal pleiotropy (MR-Egger intercept = -0.13, 95%CI -0.38–0.12, *p*=0.41), but there was some evidence of heterogeneity across individual SNP estimates (Cochran’s Q 8.90, *p*=0.03) (**Supplementary Table 8**) and no evidence against bias due to correlated pleiotropy (CAUSE OR=1.00, 95%CI 0.96–1.04, *p*=1.00; shared *q*=4%, 95%CI=0–24%) (**Supplementary Figure 4**).

Among subtypes, we found strong evidence for a causal relationship between GrimAge acceleration and colon cancer (IVW OR=1.15, 95%CI 1.09–1.21, *p*=0.006). In contrast, we did not find such evidence for rectal cancer (IVW OR=1.05, 95%CI 0.97–1.13, *p*=0.24). After further stratification, the magnitude of the genetically predicted effect of GrimAge acceleration on colon cancer was the same for distal (IVW OR=1.16, 95%CI 1.03–1.29, *p*=0.01) and proximal colon cancer (IVW OR=1.16, 95%CI 0.97–1.40, *p*=0.11). Also, sex-stratified results suggest that GrimAge acceleration may influence colorectal cancer in both males (IVW OR=1.12, 95%CI 1.00–1.25, *p*=0.05) and females (IVW OR=1.14, 95%CI 1.04–1.26, *p*=0.008) (**Figure 4, Supplementary Table 9**).

These findings were further supported by evidence of a positive association between GrimAge acceleration and parental history of colorectal cancer (OR=1.06, 95%CI 1.00–1.12, *p*=0.03) (**Figure 5, Supplementary Table 10**). Additionally, LD Score regression coefficients for GrimAge acceleration and colorectal cancer were also in the expected direction (GECCO rg=0.28, *p*<0.001; UK Biobank rg=0.15, *p*=0.21; FinnGen rg=0.27, *p*=0.29) (**Supplementary Figure 3, Supplementary Table 11**).

We did not find consistent evidence of causality between other measures of epigenetic age acceleration and colorectal cancer (PhenoAge IVW OR=1.00, 95%CI 0.97–1.02, *p*=0.73; HannumAge IVW OR=1.02, 95%CI 0.97–1.08, *p*=0.37; and Intrinsic HorvathAge IVW OR=1.00, 95%CI 0.97–1.02, *p*=0.79) (**Figure 2, Supplementary Figures 1 and 5–7,** Supplementary Tables 3–6 and 9–10).

## DISCUSSION

In this comprehensive two-sample MR study of epigenetic age acceleration and multiple cancers, we found evidence to suggest that genetically predicted GrimAge acceleration may increase the risk of colorectal cancer in both males and females. Among subtypes, effects appeared to be stronger in relation to colon than rectal cancer. Our MR results also suggested that genetically predicted GrimAge acceleration may decrease the risk of prostate cancer. The protective effect appeared strongest for advanced and early onset prostate cancer. Nevertheless, we cannot rule out the possibility of bias, as the prostate cancer results did not replicate in the MR analysis using data regarding parental history of cancer, and LD Score regression analyses did not indicate a genetic correlation between the traits.

We also identified a protective effect of Intrinsic HorvathAge acceleration on lung cancer, but this did not pass multiple testing correction and was substantially attenuated in sensitivity analyses. Finally, we found no consistent evidence for other measures of epigenetic age acceleration and cancers.

Our estimates for the association between GrimAge and colorectal cancer were consistent with those reported in Dugue et al.^10^, an observational nested case-control study in the Melbourne Collaborative Cohort Study (RR=1.04 per year increase in GrimAge acceleration, 95%CI 1.01–1.07, *p*=0.02). However, our findings contrast with those highlighted in Hillary et al.^15^. The latter authors observed no evidence of an association between GrimAge acceleration and colorectal cancer after correcting for multiple testing. Nevertheless, it is possible that their analyses were underpowered, as their sample only included 63 colorectal cancer cases (0.66%). More importantly, the direction of the reported estimate is consistent with our findings and those presented in Dugue et al.^10^.

As in our study, Dugue et al.^10^ provided evidence of a weak inverse association between GrimAge acceleration and prostate cancer (RR=0.86 per 5-year increase in GrimAge acceleration, 95%CI 0.74–1.01, *p*=0.07). Nevertheless, associations in both studies seem to have been driven by different prostate cancer subtypes. Stratified results in Dugue et al.^10^ suggested that the protective effect of GrimAge acceleration is driven by non-aggressive prostate cancer, while we only found evidence of a protective effect for early onset prostate cancer and advanced prostate cancer (which both tend to be more aggressive^37^).

The weak inverse association observed in Dugue et al.^10^ could have emerged due to detection bias. Health-conscious individuals are more likely to use health services and undergo prostate cancer screening^38^ ^39^. Hence, their likelihood of being diagnosed with less severe cancers may be greater, despite them potentially also having lower GrimAge acceleration. This could lead to spurious conclusions, such as that people with higher GrimAge acceleration are less likely to develop non-aggressive prostate cancer. The inverse effect in our study could be explained by selection bias, as cases of early onset prostate cancer (who possibly have higher GrimAge acceleration) are likely to be missed if studies only recruit middle- to late-aged participants. Likewise, individuals with longer lifespans (and possibly lower GrimAge acceleration) are more likely to develop advanced prostate cancer, simply because they have not died of other illnesses (survival bias).

Observational evidence for the association between other measures of epigenetic ageing and cancer is inconclusive (the pre-existing evidence has been summarised in **Supplementary Table 12**). For instance, epigenetic clock acceleration has been positively associated with breast^8^ ^11^ ^12^ and lung cancer^5^ ^9^ ^10^ in some studies. However, Durso et al.^16^, Hillary et al.^15^ and Dugue et al.^14^ did not find strong evidence to support this. In some cases, observational evidence is stronger for some clocks than it is for others. For example, for colorectal cancer, evidence of a positive association was much stronger for second-generation clocks^10^ than for first-generation clocks^14^ ^16^. For prostate cancer, evidence of an association was only found for GrimAge^10^ ^14^, as in our study. To date, the association between epigenetic age acceleration and ovarian cancer has not been explored observationally. Although our findings were less susceptible to biases that often influence observational research, they still provide no compelling evidence of a causal association between several measures of epigenetic clock acceleration and cancer.

This MR study had several strengths. For instance, we pooled results from multiple sources using fixed effect meta-analysis to improve the precision of the estimates presented in McCartney et al^20^. We also conducted extra sensitivity analyses, such as single-SNP and CAUSE analyses, to assess the validity of the MR assumptions. Moreover, we performed subtype-specific MR analyses and sought to replicate our results using UK Biobank GWAS data on parental history of cancer and LD Score regression. Additionally, our findings contribute to the identification of modifiable targets for future interventions aimed at reversing epigenetic ageing for the prevention of cancer. Compared to clinical trials, MR provides a cheaper, quicker, and ethical way of assessing the long-term impact of interventions on epigenetic ageing. This is especially relevant while attempts to develop interventions which reverse epigenetic ageing are still in early stages^40–43^.

The findings from this study should be interpreted in light of its limitations. We only identified four genetic instruments for GrimAge acceleration, which explained 0.47% of the variance in the trait. This could lead to two issues: low statistical power and horizontal pleiotropy. First, our GrimAge analyses were underpowered to detect ORs <1.20 for colorectal cancer and >0.84 for prostate cancer. Therefore, it is possible that our findings do not reflect a true effect (we identified ORs=1.12 and 0.93 for colorectal and prostate cancer, respectively). Similarly, our study was underpowered to detect genetically predicted effects of GrimAge acceleration on cancer subtypes and cancers with smaller sample sizes (i.e., ovarian and lung cancer). Some of our sensitivity analyses, such as the MR-Egger intercept test used to detect uncorrelated horizontal pleiotropy, also had low power, resulting in imprecise estimates. The weighted mode method may also be misleading in this context, as its use is limited in the presence of very few SNPs. Although these limitations potentially undermine the validity of our results, it is reassuring that point estimates for the genetically predicted effect of GrimAge acceleration on prostate and colorectal cancer were consistent across MR methods and study populations. However, since CAUSE analyses did not provide evidence against confounding by correlated horizontal pleiotropy, it is possible that the genetically predicted effects identified are attributed to correlated pleiotropy (whereby SNPs are associated with epigenetic age acceleration and cancer through a shared heritable factor) rather than a causal effect of GrimAge on cancer risk.

One could argue that because the results for GrimAge acceleration were inconsistent with those obtained for other measures of epigenetic age acceleration, chance and horizontal pleiotropy are more likely explanations for our findings. However, inconsistencies across epigenetic ageing measures do not necessarily invalidate our results. They may simply reflect differences in how clocks were trained (i.e., they were trained on different outcomes, tissues, and populations). Different clocks may capture information on distinct underlying biological ageing mechanisms^2^. For example, GrimAge was trained on mortality and smoking (factors which are closely related to cancer risk), which may explain why it outperforms other measures of epigenetic ageing in predicting time-to-cancer^6^. Although little is known about the underlying mechanisms, GrimAge may plausibly influence cancer risk through its effect on hormonal, inflammatory and metabolic processes^44–46^. Furthermore, Wang et al.^47^ suggests that the disruption of epigenomic maintenance systems, reflected by either epigenetic clock acceleration or deceleration, may be involved in the process of carcinogenesis.

Although promising in terms of consistency and biological plausibility, further research is required to confirm our findings. For example, multivariable MR^48^ ^49^ could be used to disentangle the causal effects of GrimAge acceleration on cancer from shared heritable factors such as and blood cell composition. Additionally, our analyses could be replicated using other large independent cancer datasets to increase power. It would also be useful to replicate our analyses once a larger GWAS of epigenetic ageing with more genetic instruments for GrimAge acceleration is available. This would allow for a more rigorous assessment of horizontal pleiotropy and may be used to assess clustering of genetic variants to reveal distinct biological mechanisms underlying the effects^50^.

The selection of “super controls” (e.g., in UK Biobank, FinnGen and GECCO), with no other cancers, related lesions (i.e., benign, in situ, uncertain or unspecified behaviour neoplasms) or reported family history of cancer, could have inflated cancer GWAS effect sizes (and our MR estimates), because “super controls” are healthier than the general population and are less likely to be genetically predisposed to develop cancer.

Another limitation is that we did not have access to individual level data. Therefore, we were unable to stratify the analyses by potential effect modifiers, such as sex, smoking and menopausal status. Moreover, we did not have sex-specific instruments for sex-specific cancers. However, it is unlikely that the genetic architecture of epigenetic clock acceleration differs across sexes, as DNAm levels at individual clock CpGs are highly correlated between males and females^51^ ^52^.

Finally, to reduce bias due to population stratification, this study was conducted using data from participants of European ancestry only. The GWAS data used for the analyses had been adjusted for the top genetic principal components for the same reason. Assortative mating is unlikely to be a problem in the context of this study because we would not expect people to select partners based on their epigenetic age acceleration. Despite this, confounding due to population stratification and assortative mating cannot be ruled out completely, as it is not possible to test the second MR assumption (i.e., independence assumption). Furthermore, more research is required to see if our results could translate to other ancestries.

In conclusion, our findings suggest that genetically predicted GrimAge acceleration may increase the risk of colorectal cancer. There is more limited evidence that it may decrease the risk of prostate cancer. Findings were less consistent for other epigenetic clocks and cancers. Further work is required to investigate the potential mechanisms underlying the genetically predicted effects identified in this study.

## Data Availability

Summary statistics for epigenetic age acceleration measures of HannumAge, Intrinsic HorvathAge, PhenoAge and GrimAge were downloaded from: https://datashare.ed.ac.uk/handle/10283/3645. Summary statistics for international cancer genetic consortiums were obtained from their respective data repositories. Colorectal cancer data were obtained following the submission of a written request to the GECCO committee, which may be contacted by email at. Breast, ovarian, prostate and lung cancer data were accessed via MR-Base (http://app.mrbase.org/), which holds complete GWAS summary data from BCAC, OCAC, PRACTICAL and ILCCO. Breast cancer subtype data were obtained from BCAC and can be downloaded from: http://bcac.ccge.medschl.cam.ac.uk/bcacdata/oncoarray/oncoarray-and-combined-summary-result/gwas-summary-associations-breast-cancer-risk-2020/. Data on breast and ovarian cancer in BRCA1 and BRCA2 carriers were obtained from CIMBA and can be downloaded from: http://cimba.ccge.medschl.cam.ac.uk/oncoarray-complete-summary-results/. Prostate cancer subtype data are not publicly available through MR-Base but can be accessed upon request. These data are managed by the PRACTICAL committee, which may be contacted by email at. FinnGen data is publicly available and can be accessed here: https://www.finngen.fi/en/access_results. UK Biobank data can be accessed through the MR-Base platform. Parental history of cancer data were obtained from the UK Biobank study under application #15825 and can be accessed via an approved application to the UK Biobank (https://www.ukbiobank.ac.uk/enable-your-research/apply-for-access).

## Acknowledgements

We thank Richard Wilkinson for proofreading several versions of the manuscript. We would also like to acknowledge the participants and investigators of the FinnGen and UK Biobank studies. GWAS data on parental history of cancer were generated using the UK Biobank Resource under application number 15825. Finally, we would like to thank the BCAC, OCAC, PRACTICAL, ILCCO, GECCO and CIMBA consortiums for their contributions.

## SUPPLEMENTARY MATERIAL

**Supplement 1. STROBE-MR checklist of recommended items to address in reports of *Mendelian randomization studies*^22^**

|  | Item<br>Nº | Recommendation | Comments/page Nº |
| --- | --- | --- | --- |
| **Title and abstract** |  |  |  |
|  | 1 | Indicate Mendelian randomization (MR) as the study's design in the title and/or the abstract if that is a main purpose of the study. | 'Mendelian randomization' in title and abstract |
| **Introduction** |  |  |  |
| Background | 2 | Explain the scientific background and rationale for the reported study. Is a potential causal relationship between exposure and outcome plausible? Justify why MR is a helpful method to address the study question. | Pages 5-7 |
| Objectives | 3 | State specific objectives clearly, including pre-specified causal hypotheses (if any). State that MR is a method that, under specific assumptions, intends to estimate causal effects. | Page 7 |
| **Methods** |  |  |  |
| Study design and data sources | 4 | Present key elements of study design early in the article. Consider including a table listing sources of data for all phases of the study. For each data source contributing to the analysis, describe the following:<br>(a) Setting: Describe the study design and the underlying population, if possible. Describe the setting, locations, and relevant dates, including periods of recruitment, exposure, follow-up, and data collection, when available.<br>(b) Participants: Give the eligibility criteria, and the sources and methods of selection of participants. Report the sample size, and whether any power or sample size calculations were carried out prior to the main analysis.<br>(c) Explain how the analysed sample size was arrived at.<br>(d) Describe measurement, quality and selection of genetic variants.<br>(e) For each exposure, outcome and other relevant variables, describe methods of assessment and diagnostic criteria for diseases.<br>(f) Provide details of ethics committee approval and participant informed consent, if relevant. | Pages 8-10, Supplementary methods, Table 1, Figure 1 |
| Assumptions | 5 | Explicitly state the three core IV assumptions for the main analysis (relevance, independence, and exclusion restriction) as well assumptions for any additional or sensitivity analysis. | Page 11, Supplementary methods |
| Statistical methods: main analysis | 6 | Describe statistical methods and statistics used.<br>(a) Describe how quantitative variables were handled in the analyses (i.e., scale, units, model).<br>(b) Describe how genetic variants were handled in the analyses and, if applicable, how their weights were selected.<br>(c) Describe the MR estimator (e.g. two-stage least squares, Wald ratio) and related statistics. Detail the included covariates and, in case of two-sample MR, whether the same covariate set was used for adjustment in the two samples.<br>(d) Explain how missing data were addressed.<br>(e) If applicable, say how multiple testing was addressed. | Page 11, Supplementary methods |
| Assessment of assumptions | 7 | Describe any methods or prior knowledge used to assess the assumptions or justify their validity. | Pages 11-13, Supplementary methods |
| Sensitivity analyses | 8 | Describe any sensitivity analyses or additional analyses performed (e.g. comparison of effect estimates from different approaches, independent replication, bias analytic techniques, validation of instruments, simulations). | Pages 11-13, Supplementary methods |
| Software and pre-registration | 9 | (a) Name statistical software and package(s), including version and settings used.<br>(b) State whether the study protocol and details were pre-registered (as well as when and where). | Pages 13-14, Supplementary methods |
| **Results** |  |  |  |
| Descriptive data | 10 | (a) Report the numbers of individuals at each stage of included studies and reasons for exclusion. Consider use of a flow diagram.<br>(b) Report summary statistics for phenotypic exposure(s), outcome(s) and other relevant variables (e.g. means, SDs, proportions).<br>(c) If the data sources include meta-analyses of previous studies, provide the assessments of heterogeneity across these studies.<br>(d) For two-sample MR:<br>i. Provide justification of the similarity of the genetic variant-exposure associations between the exposure and outcome samples.<br>ii. Provide information on the number of individuals who overlap between the exposure and outcome studies. | Not applicable, as we used summary statistics from previously published genome-wide association studies. We cite these accordingly. Furthermore, there is no sample overlap between the exposure and outcome studies. |
| Main results | 11 | (a) Report the associations between genetic variant and exposure, and between genetic variant and outcome, preferably on an interpretable scale.<br>(b) Report MR estimates of the relationship between exposure and outcome, and the measures of uncertainty from the MR analysis on an interpretable scale, such as odds ratio or relative risk per SD difference.<br>(c) If relevant, consider translating estimates of relative risk into absolute risk for a meaningful time-period.<br>(d) Consider plots to visualize results (e.g. forest plot, scatterplot of associations between genetic variants and outcome versus between genetic variants and exposure). | Pages 16-20, Figure 2, Supplementary figures and tables |
| Assessment of validity | 12 | (a) Report the assessment of the validity of the assumptions.<br>(b) Report any additional statistics (e.g., assessments of heterogeneity across genetic variants, such as $I^2$, Q statistic or E-value). | Pages 16-20 and Figure 3, Supplementary figures and tables |
| Sensitivity and additional analyses | 13 | (a) Report any sensitivity analyses to assess the robustness of the main results to violations of the assumptions.<br>(b) Report results from other sensitivity analyses or additional analyses. | Pages 16-20 and Figures 3-5, Supplementary figures and tables |
|  |  | (c) Report any assessment of direction of causal relationship (e.g., bidirectional MR). |  |
|  |  | (d) When relevant, report and compare with estimates from non-MR analyses. |  |
|  |  | (e) Consider any additional plots to visualize results (e.g., leave-one-out analyses). |  |
| **Discussion** |  |  |  |
| Key results | 14 | Summarize key results with reference to study objectives. | Page 25 |
| Limitations | 15 | Discuss limitations of the study, taking into account the validity of the IV assumptions, other sources of potential bias, and imprecision. Discuss both direction and magnitude of any potential bias, and any efforts to address them. | Pages 25-31 |
| Interpretation | 16 | (a) Meaning: Give a cautious overall interpretation of results in the context of their limitations and in comparison with other studies.<br>(b) Mechanisms: Discuss underlying biological mechanisms that could drive a potential causal relationship between the investigated exposure and the outcome, and whether the gene-environment equivalence assumption is reasonable. Use causal language carefully, clarifying that IV estimates may provide causal effects only under certain assumptions.<br>(c) Clinical relevance: Discuss whether the results have clinical or public policy relevance, and to what extent they inform effect sizes of possible interventions. | Pages 25-31 |
| Generalisability | 17 | Discuss the generalizability of the study results (a) to other populations (i.e. external validity), (b) across other exposure periods/timings, and (c) across other levels of exposure. | Page 30 |
| **Other information** |  |  |  |
| Funding | 18 | Describe sources of funding and the role of funders in the present study and, if applicable, sources of funding for the databases and original study or studies on which the present study is based. | Abstract |
| Data and data sharing | 19 | Provide the data used to perform all analyses or report where and how the data can be accessed, and reference these sources in the article. Provide the statistical code needed to reproduce the results in the article, or report whether the code is publicly accessible and if so, where. | Supplementary methods |
| Conflicts of interest | 20 | All authors should declare all potential conflicts of interest. | Page 31 |

Adapted from: BMJ 2021;375:n2233 http://dx.doi.org/10.1136/bmj.n2233

**Supplement 2. PRACTICAL consortia authorship and acknowledgements PIs from the PRACTICAL (http://practical.icr.ac.uk/) consortia:**

Rosalind A. Eeles^1,2^, Christopher A. Haiman^3^, Zsofia Kote-Jarai^1^, Fredrick R. Schumacher^4,5^, Sara Benlloch^6,1^, Ali Amin Al Olama^6,7^, Kenneth R. Muir^8^, Sonja I. Berndt^9^, David V. Conti^3^, Fredrik Wiklund^10^, Stephen Chanock^9^, Ying Wang^11^, Catherine M. Tangen^12^, Jyotsna Batra^13, 14^, Judith A. Clements^13, 14^, APCB BioResource (Australian Prostate Cancer BioResource)^14, 15^, Henrik Grönberg^10^, Nora Pashayan^16,17^, Johanna Schleutker^18,19^, Demetrius Albanes^9^, Stephanie Weinstein^9^, Alicja Wolk^20^, Catharine M. L. West^21^, Lorelei A. Mucci^22^, Géraldine Cancel-Tassin^23,24^, Stella Koutros^9^, Karina Dalsgaard Sørensen^25,26^, Eli Marie Grindedal^27^, David E. Neal^28,29,30^, Freddie C. Hamdy^31,32^, Jenny L. Donovan^33^, Ruth C. Travis^34^, Robert J. Hamilton^35,36^, Sue Ann Ingles^37^, Barry S. Rosenstein^38^, Yong-Jie Lu^39^, Graham G. Giles^40,41,42^, Robert J. MacInnis^40,41^, Adam S. Kibel^43^, Ana Vega^44,45,46^, Manolis Kogevinas^47,48,49,50^, Kathryn L. Penney^51^, Jong Y. Park^52^, Janet L. Stanford^53,54^, Cezary Cybulski^55^, Børge G. Nordestgaard^56,57^, Sune F. Nielsen^56,57^, Hermann Brenner^58,59,60^, Christiane Maier^61^, Jeri Kim^62^, Esther M. John^63^, Manuel R. Teixeira^64,65,66^, Susan L. Neuhausen^67^, Kim De Ruyck^68^, Azad Razack^69^, Lisa F. Newcomb^53,70^, Davor Lessel^71^, Radka Kaneva^72^, Nawaid Usmani^73,74^, Frank Claessens^75^, Paul A. Townsend^76,77^, Jose Esteban Castelao^78^, Monique J. Roobol^79^, Florence Menegaux^80^, Kay-Tee Khaw^81^, Lisa Cannon-Albright^82,83^, Hardev Pandha^77^, Stephen N. Thibodeau^84^, David J. Hunter^85^, Peter Kraft^86^, William J. Blot^87,88^, Elio Riboli^89^

^1^The Institute of Cancer Research, London, SM2 5NG, UK

^2^Royal Marsden NHS Foundation Trust, London, SW3 6JJ, UK

^3^Center for Genetic Epidemiology, Department of Preventive Medicine, Keck School of Medicine, University of Southern California/Norris Comprehensive Cancer Center, Los Angeles, CA 90015, USA

^4^Department of Population and Quantitative Health Sciences, Case Western Reserve University, Cleveland, OH 44106-7219, USA

^5^Seidman Cancer Center, University Hospitals, Cleveland, OH 44106, USA.

^6^Centre for Cancer Genetic Epidemiology, Department of Public Health and Primary Care, University of Cambridge, Strangeways Research Laboratory, Cambridge CB1 8RN, UK

^7^University of Cambridge, Department of Clinical Neurosciences, Stroke Research Group, R3, Box 83, Cambridge Biomedical Campus, Cambridge CB2 0QQ, UK

^8^Division of Population Health, Health Services Research and Primary Care, University of Manchester, Oxford Road, Manchester, M13 9PL, UK

^9^Division of Cancer Epidemiology and Genetics, National Cancer Institute, NIH, Bethesda, Maryland, 20892, USA

^10^Department of Medical Epidemiology and Biostatistics, Karolinska Institute, SE-171 77 Stockholm, Sweden

^11^Department of Population Science, American Cancer Society, 250 Williams Street, Atlanta, GA 30303, USA

^12^SWOG Statistical Center, Fred Hutchinson Cancer Research Center, Seattle, WA 98109, USA

^13^Australian Prostate Cancer Research Centre-Qld, Institute of Health and Biomedical Innovation and School of Biomedical Sciences, Queensland University of Technology, Brisbane QLD 4059, Australia

^14^Translational Research Institute, Brisbane, Queensland 4102, Australia

^15^Australian Prostate Cancer Research Centre-Qld, Queensland University of Technology, Brisbane; Prostate Cancer Research Program, Monash University, Melbourne; Dame Roma Mitchell Cancer Centre, University of Adelaide, Adelaide; Chris O’Brien Lifehouse

^16^Department of Applied Health Research, University College London, London, WC1E 7HB, UK

^17^Centre for Cancer Genetic Epidemiology, Department of Oncology, University of Cambridge, Strangeways Laboratory, Worts Causeway, Cambridge, CB1 8RN, UK ^18^Institute of Biomedicine, University of Turku, Finland

^19^Department of Medical Genetics, Genomics, Laboratory Division, Turku University Hospital, PO Box 52, 20521 Turku, Finland

^20^Department of Surgical Sciences, Uppsala University, 75185 Uppsala, Sweden

^21^Division of Cancer Sciences, University of Manchester, Manchester Academic Health Science Centre, Radiotherapy Related Research, The Christie Hospital NHS Foundation Trust, Manchester, M13 9PL UK

^22^Department of Epidemiology, Harvard T. H. Chan School of Public Health, Boston, MA 02115, USA

^23^CeRePP, Tenon Hospital, F-75020 Paris, France.

^24^Sorbonne Universite, GRC n°5 , AP-HP, Tenon Hospital, 4 rue de la Chine, F-75020 Paris, France

^25^Department of Molecular Medicine, Aarhus University Hospital, Palle Juul-Jensen Boulevard 99, 8200 Aarhus N, Denmark

^26^Department of Clinical Medicine, Aarhus University, DK-8200 Aarhus N

^27^Department of Medical Genetics, Oslo University Hospital, 0424 Oslo, Norway

^28^Nuffield Department of Surgical Sciences, University of Oxford, Room 6603, Level 6, John Radcliffe Hospital, Headley Way, Headington, Oxford, OX3 9DU, UK

^29^University of Cambridge, Department of Oncology, Box 279, Addenbrooke’s Hospital, Hills Road, Cambridge CB2 0QQ, UK

^30^Cancer Research UK, Cambridge Research Institute, Li Ka Shing Centre, Cambridge, CB2 0RE, UK

^31^Nuffield Department of Surgical Sciences, University of Oxford, Oxford, OX1 2JD, UK

^32^Faculty of Medical Science, University of Oxford, John Radcliffe Hospital, Oxford, UK

^33^Population Health Sciences, Bristol Medical School, University of Bristol, BS8 2PS, UK

^34^Cancer Epidemiology Unit, Nuffield Department of Population Health, University of Oxford, Oxford, OX3 7LF, UK

^35^Dept. of Surgical Oncology, Princess Margaret Cancer Centre, Toronto ON M5G 2M9, Canada

^36^Dept. of Surgery (Urology), University of Toronto, Canada

^37^Department of Preventive Medicine, Keck School of Medicine, University of Southern California/Norris Comprehensive Cancer Center, Los Angeles, CA 90015, USA

^38^Department of Radiation Oncology and Department of Genetics and Genomic Sciences, Box 1236, Icahn School of Medicine at Mount Sinai, One Gustave L. Levy Place, New York, NY 10029, USA

^39^Centre for Cancer Biomarker and Biotherapeutics, Barts Cancer Institute, Queen Mary University of London, John Vane Science Centre, Charterhouse Square, London, EC1M 6BQ, UK

^40^Cancer Epidemiology Division, Cancer Council Victoria, 615 St Kilda Road, Melbourne, VIC 3004, Australia

^41^Centre for Epidemiology and Biostatistics, Melbourne School of Population and Global Health, The University of Melbourne, Grattan Street, Parkville, VIC 3010, Australia

^42^Precision Medicine, School of Clinical Sciences at Monash Health, Monash University, Clayton, Victoria 3168, Australia

^43^Division of Urologic Surgery, Brigham and Womens Hospital, 75 Francis Street, Boston, MA 02115, USA

^44^Fundación Pública Galega Medicina Xenómica, Santiago de Compostela, 15706, Spain

^45^Instituto de Investigación Sanitaria de Santiago de Compostela, Santiago De Compostela, 15706, Spain.

^46^Centro de Investigación en Red de Enfermedades Raras (CIBERER), Spain

^47^ISGlobal, Barcelona, Spain

^48^IMIM (Hospital del Mar Medical Research Institute), Barcelona, Spain

^49^Universitat Pompeu Fabra (UPF), Barcelona, Spain

^50^CIBER Epidemiología y Salud Pública (CIBERESP), 28029 Madrid, Spain

^51^Channing Division of Network Medicine, Department of Medicine, Brigham and Women’s Hospital/Harvard Medical School, Boston, MA 02115, USA

^52^Department of Cancer Epidemiology, Moffitt Cancer Center, 12902 Magnolia Drive, Tampa, FL 33612, USA

^53^Division of Public Health Sciences, Fred Hutchinson Cancer Research Center, Seattle, Washington, 98109-1024, USA

^54^Department of Epidemiology, School of Public Health, University of Washington, Seattle, Washington 98195, USA

^55^International Hereditary Cancer Center, Department of Genetics and Pathology, Pomeranian Medical University, 70-115 Szczecin, Poland

^56^Faculty of Health and Medical Sciences, University of Copenhagen, 2200 Copenhagen, Denmark

^57^Department of Clinical Biochemistry, Herlev and Gentofte Hospital, Copenhagen University Hospital, Herlev, 2200 Copenhagen, Denmark

^58^Division of Clinical Epidemiology and Aging Research, German Cancer Research Center (DKFZ), D-69120, Heidelberg, Germany

^59^German Cancer Consortium (DKTK), German Cancer Research Center (DKFZ), D-69120 Heidelberg, Germany

^60^Division of Preventive Oncology, German Cancer Research Center (DKFZ) and National Center for Tumor Diseases (NCT), Im Neuenheimer Feld 460, 69120 Heidelberg, Germany

^61^Humangenetik Tuebingen, Paul-Ehrlich-Str 23, D-72076 Tuebingen, Germany

^62^The University of Texas M. D. Anderson Cancer Center, Department of Genitourinary Medical Oncology, 1515 Holcombe Blvd., Houston, TX 77030, USA

^63^Departments of Epidemiology & Population Health and of Medicine, Division of Oncology, Stanford Cancer Institute, Stanford University School of Medicine, Stanford, CA 94304 USA

^64^Department of Genetics, Portuguese Oncology Institute of Porto (IPO-Porto), 4200-072 Porto, Portugal

^65^Biomedical Sciences Institute (ICBAS), University of Porto, 4050-313 Porto, Portugal

^66^Cancer Genetics Group, IPO-Porto Research Center (CI-IPOP), Portuguese Oncology Institute of Porto (IPO-Porto), 4200-072 Porto, Portugal

^67^Department of Population Sciences, Beckman Research Institute of the City of Hope, 1500 East Duarte Road, Duarte, CA 91010, 626-256-HOPE (4673)

^68^Ghent University, Faculty of Medicine and Health Sciences, Basic Medical Sciences, Proeftuinstraat 86, B-9000 Gent

^69^Department of Surgery, Faculty of Medicine, University of Malaya, 50603 Kuala Lumpur, Malaysia

^70^Department of Urology, University of Washington, 1959 NE Pacific Street, Box 356510, Seattle, WA 98195, USA

^71^Institute of Human Genetics, University Medical Center Hamburg-Eppendorf, D-20246 Hamburg, Germany

^72^Molecular Medicine Center, Department of Medical Chemistry and Biochemistry, Medical University of Sofia, Sofia, 2 Zdrave Str., 1431 Sofia, Bulgaria

^73^Department of Oncology, Cross Cancer Institute, University of Alberta, 11560 University Avenue, Edmonton, Alberta, Canada T6G 1Z2

^74^Division of Radiation Oncology, Cross Cancer Institute, 11560 University Avenue, Edmonton, Alberta, Canada T6G 1Z2

^75^Molecular Endocrinology Laboratory, Department of Cellular and Molecular Medicine, KU Leuven, BE-3000, Belgium

^76^Division of Cancer Sciences, Manchester Cancer Research Centre, Faculty of Biology, Medicine and Health, Manchester Academic Health Science Centre, NIHR Manchester Biomedical Research Centre, Health Innovation Manchester, Univeristy of Manchester, M13 9WL

^77^The University of Surrey, Guildford, Surrey, GU2 7XH, UK

^78^Genetic Oncology Unit, CHUVI Hospital, Complexo Hospitalario Universitario de Vigo, Instituto de Investigación Biomédica Galicia Sur (IISGS), 36204, Vigo (Pontevedra), Spain

^79^Department of Urology, Erasmus University Medical Center, 3015 CE Rotterdam, The Netherlands

^80^"Exposome and Heredity", CESP (UMR 1018), Faculté de Médecine, Université Paris-Saclay, Inserm, Gustave Roussy, Villejuif

^81^Clinical Gerontology Unit, University of Cambridge, Cambridge, CB2 2QQ, UK

^82^Division of Epidemiology, Department of Internal Medicine, University of Utah School of Medicine, Salt Lake City, Utah 84132, USA

^83^George E. Wahlen Department of Veterans Affairs Medical Center, Salt Lake City, Utah 84148, USA

^84^Department of Laboratory Medicine and Pathology, Mayo Clinic, Rochester, MN 55905, USA

^85^Nuffield Department of Population Health, University of Oxford, United Kingdom

^86^Program in Genetic Epidemiology and Statistical Genetics, Department of Epidemiology, Harvard School of Public Health, Boston, MA, USA

^87^Division of Epidemiology, Department of Medicine, Vanderbilt University Medical Center, 2525 West End Avenue, Suite 800, Nashville, TN 37232 USA.

^88^International Epidemiology Institute, Rockville, MD 20850, USA

^89^Department of Epidemiology and Biostatistics, School of Public Health, Imperial College London, SW7 2AZ, UK

## CRUK and PRACTICAL consortium Funding Acknowledgements

This work was supported by the Canadian Institutes of Health Research, European Commission’s Seventh Framework Programme grant agreement n° 223175 (HEALTH-F2-2009-223175), Cancer Research UK Grants C5047/A7357, C1287/A10118, C1287/A16563, C5047/A3354, C5047/A10692, C16913/A6135, and The National Institute of Health (NIH) Cancer Post-Cancer GWAS initiative grant: No. 1 U19 CA 148537-01 (the GAME-ON initiative).

We would also like to thank the following for funding support: The Institute of Cancer Research and The Everyman Campaign, The Prostate Cancer Research Foundation, Prostate Research Campaign UK (now PCUK), The Orchid Cancer Appeal, Rosetrees Trust, The National Cancer Research Network UK, The National Cancer Research Institute (NCRI) UK. We are grateful for support of NIHR funding to the NIHR Biomedical Research Centre at The Institute of Cancer Research, The Royal Marsden NHS Foundation Trust, and Manchester NIHR Biomedical Research Centre. The Prostate Cancer Program of Cancer Council Victoria also acknowledge grant support from The National Health and Medical Research Council, Australia (126402, 209057, 251533, , 396414, 450104, 504700, 504702, 504715, 623204, 940394, 614296,), VicHealth, Cancer Council Victoria, The Prostate Cancer Foundation of Australia, The Whitten Foundation, PricewaterhouseCoopers, and Tattersall’s. EAO, DMK, and EMK acknowledge the Intramural Program of the National Human Genome Research Institute for their support.

Genotyping of the OncoArray was funded by the US National Institutes of Health (NIH) [U19 CA 148537 for ELucidating Loci Involved in Prostate cancer SuscEptibility (ELLIPSE) project and X01HG007492 to the Center for Inherited Disease Research (CIDR) under contract number HHSN268201200008I]. Additional analytic support was provided by NIH NCI U01 CA188392 (PI: Schumacher).

Research reported in this publication also received support from the National Cancer Institute of the National Institutes of Health under Award Numbers U10 CA37429 (CD Blanke), and UM1 CA182883 (CM Tangen/IM Thompson). The content is solely the responsibility of the authors and does not necessarily represent the official views of the National Institutes of Health.

Funding for the iCOGS infrastructure came from: the European Community’s Seventh Framework Programme under grant agreement n° 223175 (HEALTH-F2-2009-223175) (COGS), Cancer Research UK (C1287/A10118, C1287/A 10710, C12292/A11174, C1281/A12014, C5047/A8384, C5047/A15007, C5047/A10692, C8197/A16565), the National Institutes of Health (CA128978) and Post-Cancer GWAS initiative (1U19 CA148537, 1U19 CA148065 and 1U19 CA148112 - the GAME-ON initiative), the Department of Defence (W81XWH-10-1-0341), the Canadian Institutes of Health Research (CIHR) for the CIHR Team in Familial Risks of Breast Cancer, Komen Foundation for the Cure, the Breast Cancer Research Foundation, and the Ovarian Cancer Research Fund.

## SUPPLEMENTARY METHODS

### Cancer datasets

#### UK Biobank

The UK Biobank is a large cohort study including around 500,000 individuals aged 40 to 69 years at the time of recruitment (2006–2010). The cohort has been described in detail in previous publications^1^ ^2^. In short, all participants provided written informed consent, after which baseline data were collected using sociodemographic, lifestyle and health-related questionnaires, physical and cognitive assessments, and biological samples. Participants’ data were linked to their health records for longitudinal follow-up. The study obtained ethical approval from the National Information Governance Board for Health and Social Care and the North-West Multicenter Research Ethics Committee (Ref: 11/NW/0382).

Cancer cases (diagnosed prior or after enrolment) were obtained from the UK Cancer Registry (updated to April 2019). They were then coded according to the ninth and tenth editions of the International Classification of Diseases (ICD-9 and ICD-10, respectively) as follows: breast (ICD-9: 174; ICD-10: C50), ovarian (ICD-9: 183; ICD-10: C56), prostate (ICD-9: 185; ICD-10: C61), lung (ICD-9: 162; ICD-10: C34) and colorectal cancer (ICD-9: 153; ICD-10: C18-C20). Controls excluded individuals with any type of cancer (self-reported and/or recorded in cancer registry), as well as those with benign, in situ, uncertain or unspecified behaviour neoplasms (ICD-9: 210-239; ICD-10: D00-D49).

Sample-level quality control (QC) involved removing any individuals who had non-white British genetic ancestry, sex chromosome aneuploidies, who withdrew consent from the UK Biobank study and who were closely related to other participants. Variant-level QC consisted in imputing SNPs using the Haplotype Reference Consortium (HRC) and restricting SNPs to a minor allele frequency (MAF) >0.1%, a genotyping rate > 0.015 and a Hardy-Weinberg Equilibrium (HWE) *p* >1x10^-4^. LD pruning was performed to an r^2^ cutoff of 0.1 using PLINK v2^3^. In order to reduce false positive signals, SNPs were removed when MAF was below our expectations (we would expect at least 25 minor alleles in cases), as recommended in http://www.nealelab.is/blog/2017/9/11/details-and-considerations-of-the-uk-biobank-gwas.

The GWAS analysis in the UK Biobank consisted of 13,879 cases and 198,523 controls for breast cancer, 1,218 cases and 198,523 controls for ovarian cancer, 9,132 cases and 173,493 controls for prostate cancer, 2,671 cases and 372,016 controls for lung cancer and 5,657 cases and 372,016 controls for colorectal cancer. It was performed using BOLT-LMM v2.3.5^4^ ^5^, adjusting for sex and genotyping chip. BOLT-LMM uses a linear mixed model to account for population stratification and cryptic relatedness in the UK Biobank. Lung cancer associations were estimated twice, once adjusting for genotyping chip and once without. For sex-specific cancers, analyses were limited to individuals of the pertinent sex (only females were used for breast and ovarian cancers, whereas only males were used for prostate cancer). Beta coefficients and their corresponding standard errors were finally transformed to log odds ratios (ORs)^5^.

We also performed a GWAS analysis of parental history of cancer reported by UK Biobank participants (i.e., breast, prostate, lung and bowel cancer) using BOLT-LMM software v2.3.5^4^. Age and sex were included as covariates in the model as before. For sex-specific cancers, analyses were restricted to individuals of the relevant sex (i.e., maternal history only for breast cancer and paternal history only for prostate cancer). We obtained 35,356 breast cancer cases and 206,992 controls, in addition to 31,527 prostate cancer cases and 160,579 controls. For other cancers, we combined maternal and paternal history of cancer, thus obtaining a total of 51,073 lung cancer cases and 404,606 controls, as well as 45,213 bowel cancer cases and 412,429 controls. GWAS of these outcomes have previously provided strong concordance with those based on hospital records^6^. They have also provided consistent results in MR^7^.

#### FinnGen

The FinnGen R5 release includes data on 218,792 individuals of Finnish ancestry, obtained from Finnish biobanks and digital health registry records^8^. Complete study details are available elsewhere (https://www.finngen.fi/en). In brief, samples were excluded for the following reasons: ambiguous gender, genotype missingness >5%, heterozygosity +-4 s.d. and non-Finnish ancestry. SNPs were genotyped using Illumina and Affymetrix arrays.

Variants were excluded for the following reasons: missingness >2%, HWE *p* <1x10-6 and minor allele count <3. Genotypes were imputed using the Finnish SISu v3 reference panel. The GWAS analysis was conducted using SAIGE v0.36.3.2, a mixed model logistic regression R/C++ package. Sex, age, genotyping batch and the first 10 genetically derived principal components were included as covariates in the analysis. We used FinnGen R5 release data on breast (8,401 cases and 99,321 controls), ovarian (719 cases and 99,321 controls), prostate (6,311 cases and 74,685 controls), lung (1,681 cases and 173,933 controls) and colorectal cancer (3,022 cases and 174,006 controls). We used the “EXALLC” cancer variables, which excluded other cancers from controls.

#### Breast Cancer Association Consortium

The GWAS summary data for breast cancer were obtained from a Breast Cancer Association Consortium (BCAC) meta-analysis performed by Michailidou et al.^9^. This included 122,977 cases and 105,974 controls (69,501 cases of ER+ and 21,468 of ER- breast cancer). All studies that contributed to this meta-analysis have been fully detailed in previous publications^9–11^.

In sum, samples were excluded if they had a low call rate (<95%), abnormally high or low heterozygosity (4.89 s.d. from the mean), <80% European ancestry, probable duplicates and/or close relatives within and across studies. Genetic variants were genotyped using the Illumina OncoArray and iCOGS arrays and genotypes were imputed using the 1000 Genomes Project Phase 3 reference panel.

We also obtained summary data for breast cancer subtypes from a BCAC GWAS meta-analysis by Zhang et al.^12^. The study comprised data on luminal A-like (7,325 cases), luminal B-like (1,682 cases), luminal B/HER2-negative-like (1,779 cases), HER2-enriched-like (718 cases) and triple-negative (2,006 cases) invasive breast cancer subtypes and 20,815 controls. The details of the study can be found in the publication. In brief, the analyses excluded cases of carcinoma in situ, cases missing data on tumour characteristics and cases for which there were no controls available in their respective countries. Participants were also excluded if age at diagnosis/enrolment was missing. Genotypes were obtained using OncoArray and iCOGS arrays. Imputation was performed using the 1000 Genomes Project Phase 3 reference panel. OncoArray and iCOGS datasets were analysed separately and pooled using fixed-effect meta-analysis.

#### Ovarian Cancer Association Consortium

We used ovarian cancer genetic summary statistics from an Ovarian Cancer Association Consortium (OCAC) study by Phelan et al.^13^. This comprised 25,509 cases and 40,941 controls. Subtypes included high grade serous (13,037 cases), low grade serous (1,012 cases), invasive mucinous (1,417 cases), clear cell (1,366 cases) and endometrioid (2,810 cases) ovarian cancers. This study combined genotype data from OCAC and Consortium of Investigators of Modifiers of BRCA1/2 (CIMBA) genotyping projects. These have been fully described in the publication. In short, samples with >27% non-European ancestry were excluded, as were those with a genotyping call rate <95%, excessively low or high heterozygosity. Non-females and duplicates were also removed. SNPs were genotyped using several Illumina arrays (OncoArray, iSelect iCOGS, 550k, HumanOmni 2.5M, 610 Quad and 317k). Imputations were performed separately for each genotyping project using the 1000 Genomes Project v3 reference panel.

#### Consortium of Investigators of Modifiers of BRCA1/2

We also used CIMBA GWAS data for breast and ovarian cancers in BRCA1 and BRCA2 mutation carriers^13^ ^14^. The genotyping and imputation procedures that were used have been described elsewhere^13^ ^14^. In brief, samples were excluded if they were non-female, had discordant genotypes in known sample duplicates, had >19% non-European ancestry, a genotyping call rate <95% or extremely low or high heterozygosity (*p*<1x10^−6^). SNPs were genotyped using Illumina’s Oncoarray and iSelect Collaborative Oncological Gene-Environment Study (iCOGS) arrays. Imputation was performed using the 1000 Genomes Project Phase 3 reference panel.

#### Prostate Cancer Association Group to Investigate Cancer Associated Alterations in the Genome

Prostate cancer GWAS summary data were acquired from a Prostate Cancer Association Group to Investigate Cancer Associated Alterations in the Genome (PRACTICAL) study by Schumacher et al.^15^. This included 79,148 cases and 61,106 controls. It also comprised data on prostate cancer subtypes: 15,167 advanced cases vs. 58,308 healthy controls; 14,160 advanced cases vs. 62,421 non-advanced controls; 6,988 early-onset cases (age at diagnosis ≤55 years) vs. 44,256 healthy controls; 15,561 high aggressive cases vs. 9,739 low aggressive controls; and 20,658 high aggressive cases vs. 38,093 low/intermediate aggressive controls.

Prostate cancer aggressiveness was defined as follows:

- Low aggressive: tumor stage ≤T1 ***AND*** Gleason score ≤6 ***AND*** prostate-specific antigen (PSA) <10 ng/mL.

- Intermediate aggressive: tumor stage T2 ***OR*** Gleason score = 7 ***OR*** PSA 10–20 ng/mL.

- High aggressive: tumor stage T3/T4, N1 or M1 ***OR*** Gleason score ≥8 ***OR*** PSA>20 ng/mL.

- Advanced: metastatic disease ***OR*** Gleason score ≥8 ***OR*** PSA >100 ng/mL ***OR*** death due to prostate cancer.

Study details are available in the publication. In brief, individuals were excluded if they presented a call rate <95%, extreme heterozygosity (>4.9 s.d. from the mean), if they were duplicates or if they were related to other participants. Only men of European ancestry (>80%) were included in the GWAS. Studies were genotyped using Illumina (OncoArray, Human 610, 60k, Infinium HumanHap 550, iSELECT, iCOGS and Human Omni 2.5) and Affymetrix GeneChip (500k and 5.0k) genotyping arrays and SNPs were imputed to the 1000 Genomes Project Phase 3 reference panel.

#### International Lung Cancer Consortium

For lung cancer, we used genetic summary data obtained from an International Lung Cancer Consortium (ILCCO) GWAS meta-analysis of 11,348 cases and 15,861 controls by Wang et al.^16^. We also used lung cancer subtype data including 3,275 squamous cell lung carcinoma cases and 15,038 controls, as well as 3,442 lung adenocarcinoma cases and 14,894 controls. Individual studies included in the meta-analysis have been explained in prior publications^17–20^. In summary, sample QC consisted in excluding any individuals of non-European ancestry, with low call rates (<90%), or abnormally high or low heterozygosity (*p*<1x10^-4^). Duplicates and closely related individuals were also removed. Genotyping was performed using Illumina HumanHap 317k, 317k+240S, 370Duo, 550k, 610k or 1M arrays. SNPs were imputed from the 1000 Genomes Project Phase 1 v3 reference panel.

#### Genetics and Epidemiology of Colorectal Cancer Consortium

Colorectal cancer GWAS summary statistics were retrieved from a Genetics and Epidemiology of Colorectal Cancer Consortium (GECCO) GWAS meta-analysis by Huyghe et al.^21^. This comprised 58,131 cases (31,288 male and 26,843 female) and 67,347 controls (34,527 male and 32,820 female). Cases were defined as patients with colorectal cancer or advanced adenoma.

Data on colorectal cancer subtypes were obtained from another GECCO publication by Huyghe et al.^22^. This included 48,214 cases and 64,159 controls (32,002 colon, 15,706 proximal colon, 14,376 distal colon and 16,212 rectal cancer cases).

Colorectal cancer subtypes were defined as follows:

- Proximal colon cancer: any primary tumour starting in the cecum, ascending colon, hepatic flexure, or transverse colon (ICD-9: 153.4, 153.6, 153.0, or 153.1, respectively).

- Distal colon cancer: any primary tumour starting in the splenic flexure, descending colon, or sigmoid colon (ICD-9 codes: 153.7, 153.2, or 153.3, respectively)

- Colon cancer: proximal and distal colon cancer cases, in addition to colon cancer cases with unspecified site.

- Rectal cancer: any primary tumour starting in the rectum or rectosigmoid junction (ICD-9 codes: 154.1, or 154.0, respectively)

Controls excluded individuals with known history of cancer or reported family history of colorectal cancer. QC procedures have been explained in the publications^21^ ^22^. In brief, the studies excluded samples with evidence of DNA contamination, high missing genotype rates, unintentional duplicate pairs and sex discrepancies. Closely related individuals and those of non-European ancestry were also excluded. Genotyping was conducted using Illumina (300k, Oncoarray, 1M, 550k, 610k, OmniExpress, OmniExpressExome, 300/240S and custom iSelect) and Affymetrix (Axiom and 500k) arrays. Imputation was performed to the HRC reference panel.

### Sensitivity analyses

MR-Egger assumes that the association between SNPs and epigenetic age acceleration is not correlated with SNPs that affect cancer via pleiotropic pathways (Instrument Strength Independent of Direct Effect—InSIDE assumption)^23^. The weighted median method assumes that at least half of the SNPs in the analysis are valid instruments. The weighted mode approach presupposes that the most frequent association estimate is not affected by pleiotropy, meaning it must correspond to the true causal effect (ZEro Modal Pleiotropy Assumption—ZEMPA)^24^.

### Code availability

The GWAS analysis for cancers in UK Biobank was performed using BOLT-LMM v2.3.5 (http://data.broadinstitute.org/alkesgroup/BOLT-LMM/). All MR analyses and visualisations were conducted using R software v4.0.2 (https://www.r-project.org/). For cancer datasets that were not obtained from the MR-Base platform, LD proxies were identified using the “LDlinkR” v1.1.2 R package (https://github.com/CBIIT/LDlinkR). Two-sample MR analyses were conducted using the “TwoSampleMR” v0.5.6 R package (https://github.com/MRCIEU/TwoSampleMR). Meta-analyses were performed using the “meta” v4.18 R package (https://github.com/guido-s/meta/). MR-CAUSE analyses were conducted using the “cause” v1.2.0 R package (https://github.com/jean997/cause). Plots were created using the “ggforestplot” v0.1.0 R package (https://github.com/nightingalehealth/ggforestplot). LD scores were computed using the “ldsc” v1.0.1 command line tool (https://github.com/bulik/ldsc). The code used in this study is available at: https://github.com/fernandam93/epiclocks_cancer.

## Supplementary Figures

**Supplementary Figure 1.**
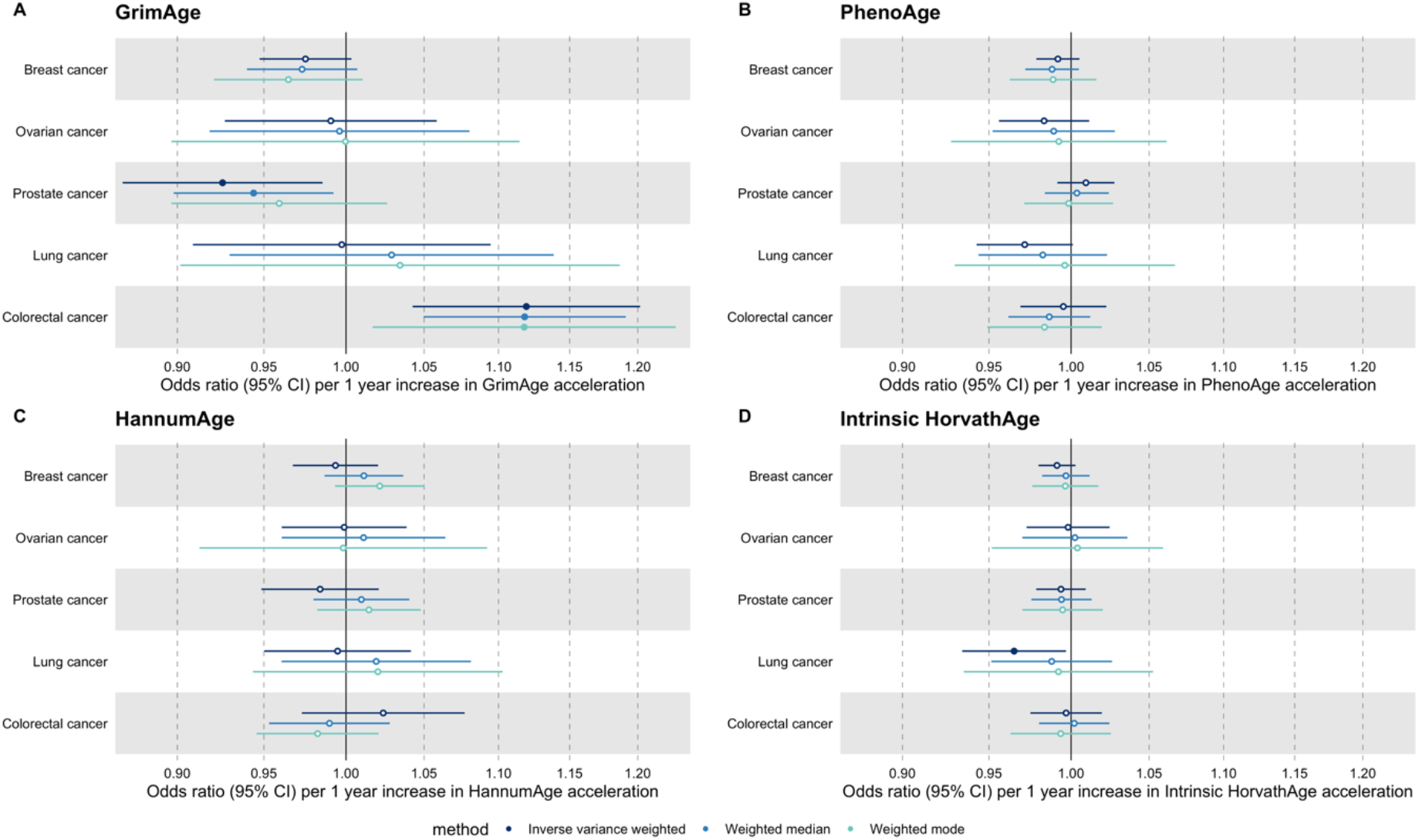
Fixed effect meta-analysis of Mendelian randomization estimates for genetically predicted effects of epigenetic age acceleration on multiple cancers. Odds ratios and 95% confidence intervals are reported per 1 year increase in (A) GrimAge acceleration, (B) PhenoAge acceleration, (C) HannumAge acceleration and (D) Intrinsic HorvathAge acceleration. Results were obtained using inverse variance weighted MR (dark blue), weighted median (sky blue) and weighted mode (turquoise) methods. All meta-analysis estimates were calculated using data from UK Biobank, FinnGen and international consortia, except for colorectal cancer estimates, which exclude UK Biobank data to avoid double counting.

**Supplementary Figure 2.**
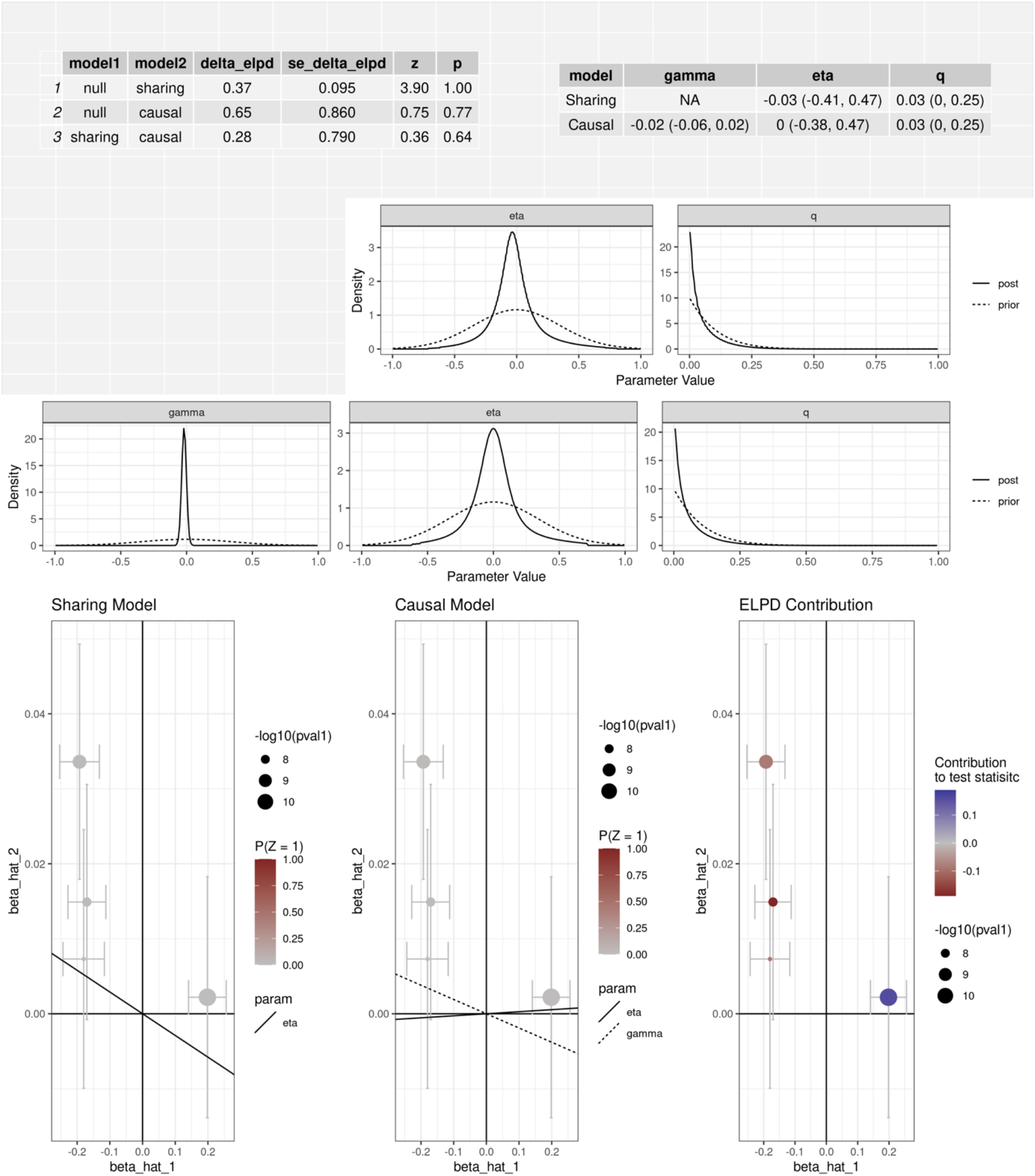
CAUSE analysis for the genetically predicted effect of GrimAge acceleration on prostate cancer in PRACTICAL. CAUSE estimates reported per 1 year increase in GrimAge acceleration. Data source: PRACTICAL, Prostate Cancer Association Group to Investigate Cancer Associated Alterations in the Genome.

**Supplementary Figure 3.**
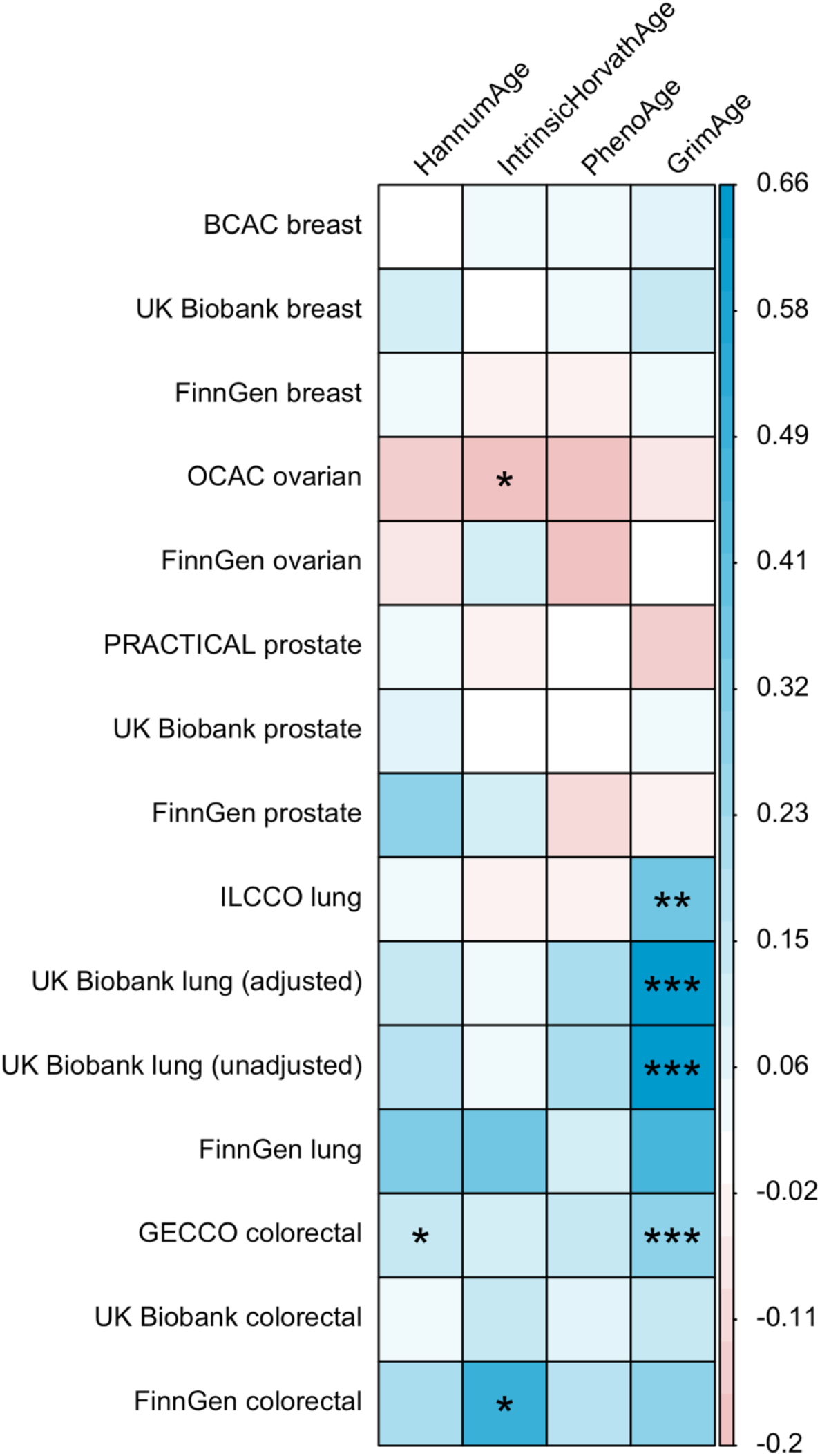
Genetic correlation estimates for epigenetic age acceleration and multiple cancers. Genetic correlation coefficients are reported per 1 year increase in epigenetic age acceleration. Results were obtained using LD Score regression. Abbreviations: BCAC, Breast Cancer Association Consortium; OCAC, Ovarian Cancer Association Consortium; PRACTICAL, Prostate Cancer Association Group to Investigate Cancer Associated Alterations in the Genome; ILCCO, International Lung Cancer Consortium; GECCO, Genetics and Epidemiology of Colorectal Cancer Consortium. For UK Biobank lung cancer results, adjusted means results have been adjusted for genotyping chip and unadjusted means results have not been adjusted for genotyping chip.

**Supplementary Figure 4.**
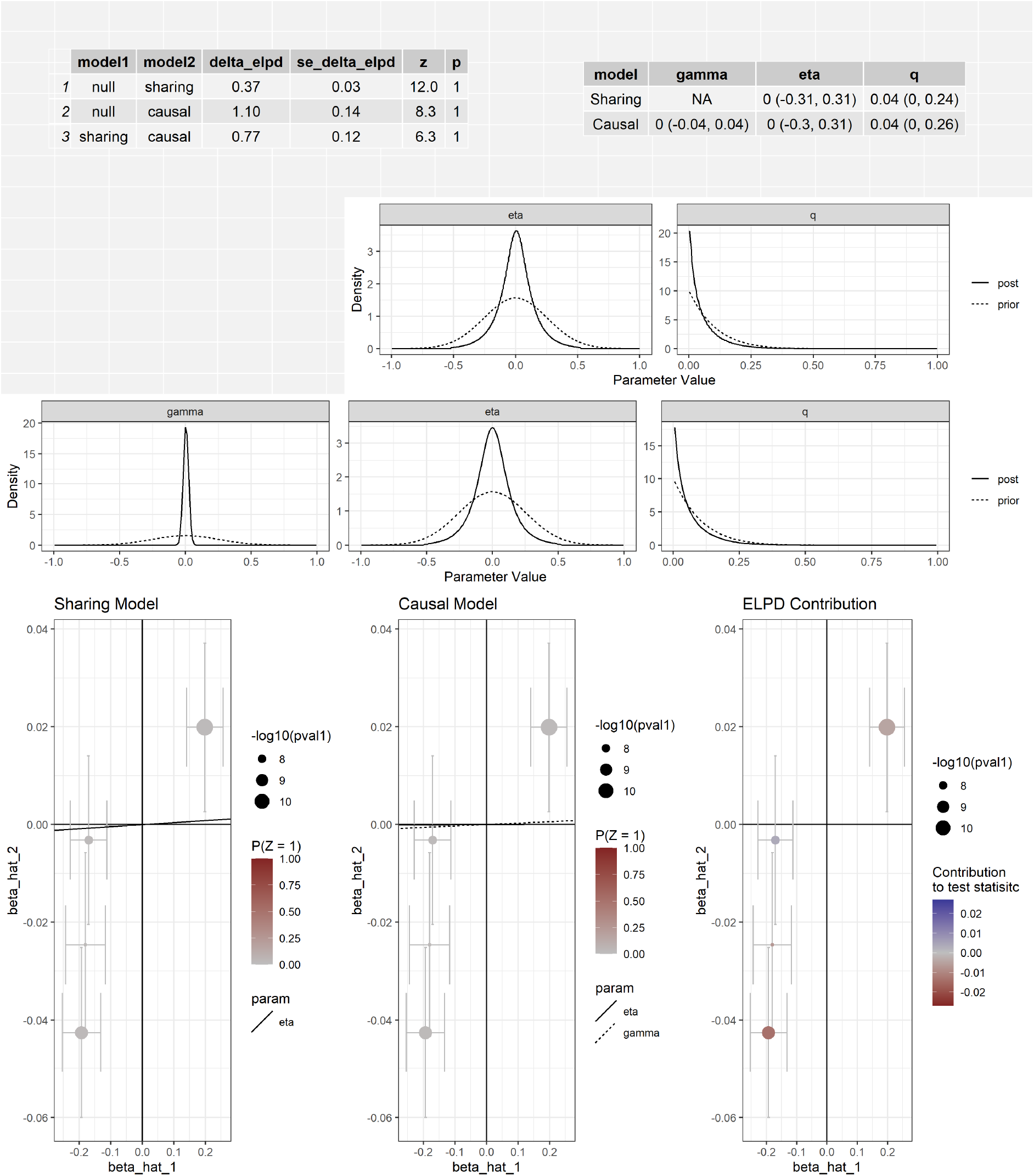
CAUSE analysis for the genetically predicted effect of GrimAge acceleration on colorectal cancer in GECCO.CAUSE estimates reported per 1 year increase in GrimAge acceleration. Data source: GECCO, Genetics and Epidemiology of Colorectal Cancer Consortium.

**Supplementary Figure 5.**
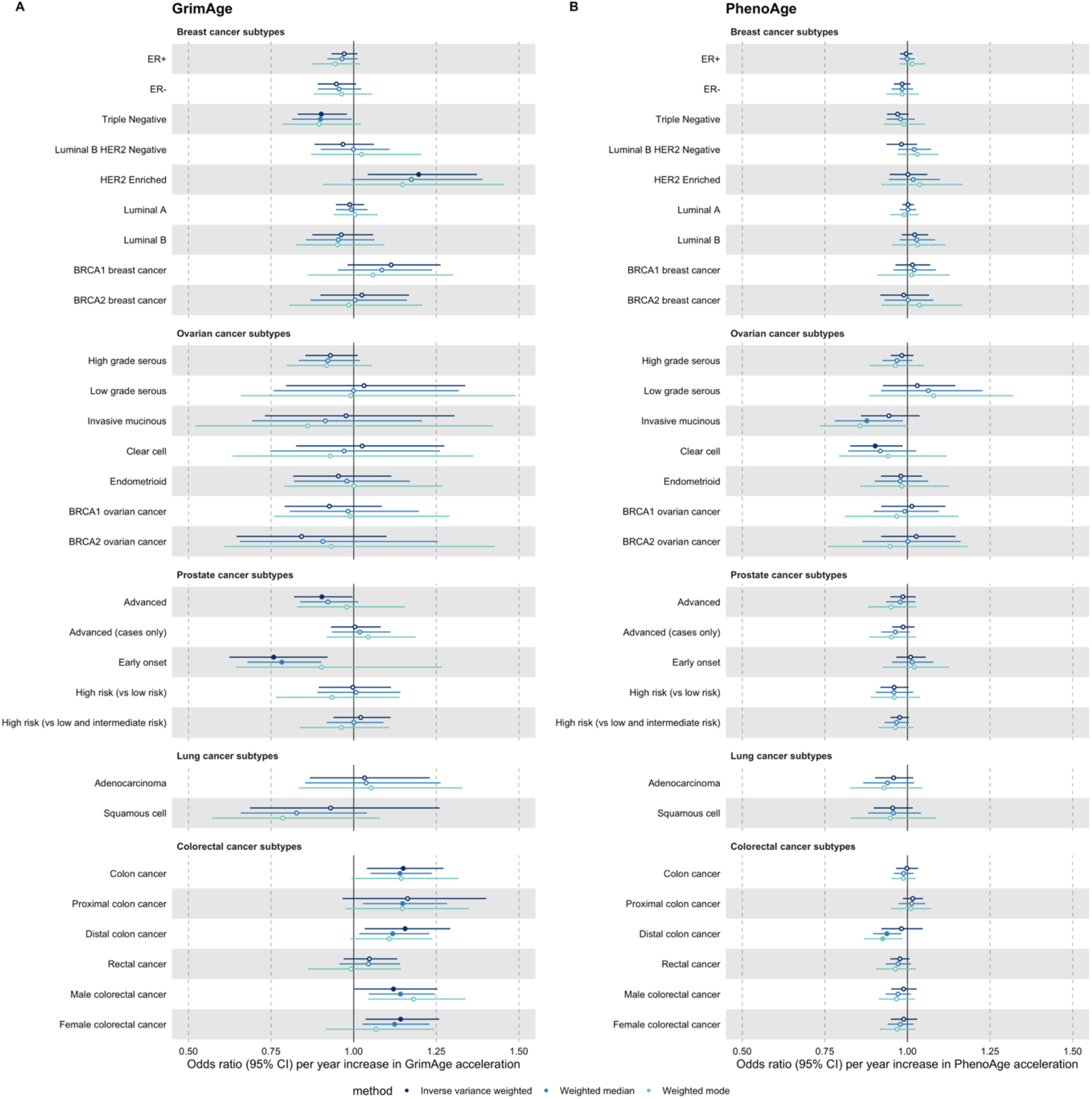
Mendelian randomization estimates for genetically predicted effects of GrimAge and PhenoAge acceleration on multiple cancer subtypes. Odds ratios and 95% confidence intervals are reported per 1 year increase in (A) GrimAge acceleration, (B) PhenoAge acceleration. GrimAge and PhenoAge acceleration were instrumented by four and 11 genetic variants, respectively. Results were obtained using inverse variance weighted MR (dark blue), weighted median (sky blue) and weighted mode (turquoise) methods. Data sources: BCAC, OCAC, CIMBA, PRACTICAL, ILCCO and GECCO.

**Supplementary Figure 6.**
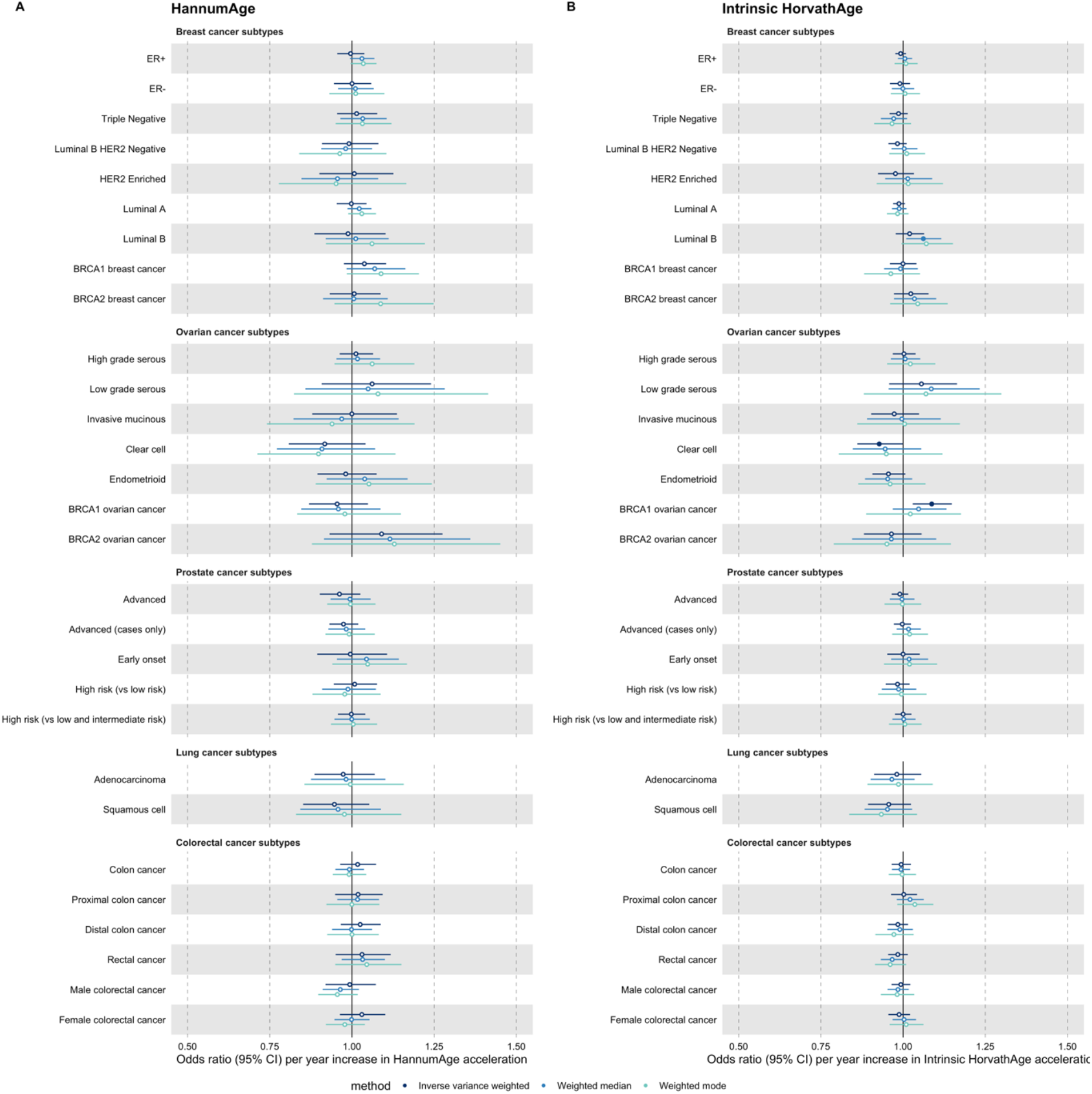
Mendelian randomization estimates for genetically predicted effects of HannumAge and Intrinsic HorvathAge acceleration on multiple cancer subtypes. Odds ratios and 95% confidence intervals are reported per 1 year increase in (A) HannumAge acceleration, (B) Intrinsic HorvathAge acceleration. HannumAge and Intrinsic HorvathAge acceleration were instrumented by nine and 24 genetic variants, respectively. Results were obtained using inverse variance weighted MR (dark blue), weighted median (sky blue) and weighted mode (turquoise) methods. Data sources: BCAC, OCAC, CIMBA, PRACTICAL, ILCCO and GECCO.

**Supplementary Figure 7.**
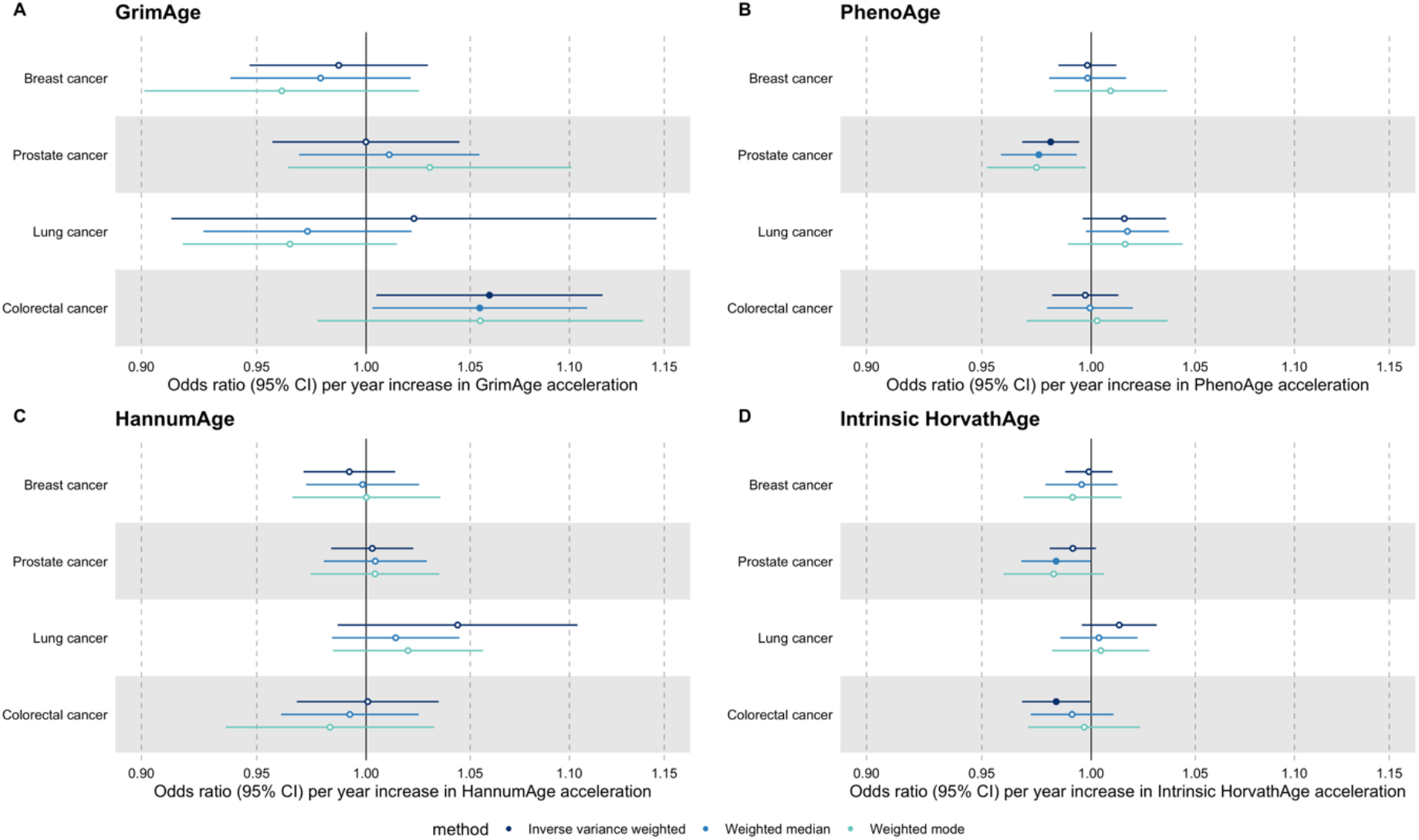
Mendelian randomization estimates for genetically predicted effects of epigenetic age acceleration on parental history of multiple cancers. Odds ratios and 95% confidence intervals are reported per 1 year increase in (A) GrimAge acceleration, (B) PhenoAge acceleration, (C) HannumAge acceleration and (D) Intrinsic HorvathAge acceleration. GrimAge, PhenoAge, HannumAge and Intrinsic HorvathAge acceleration were instrumented by 4, 11, 9 and 24 genetic variants, respectively. Results were obtained using inverse variance weighted MR (dark blue), weighted median (sky blue) and weighted mode (turquoise) methods. Data source: UK Biobank.

